# African-specific genetic loci determine iron status and risk of severe malaria and bacteremia in African children

**DOI:** 10.1101/2025.03.02.25323069

**Authors:** John Muthii Muriuki, Alexander J Mentzer, Gavin Band, Amanda Y Chong, Alex W Macharia, Reagan M Mogire, Kelvin Mokaya Abuga, Ruth Mitchell, James J Gilchrist, Emily L Webb, Francis M Ndungu, Laura M Raffield, Lynette Ekunwe, Amy R Bentley, Sodiomon B Sirima, Shabir A Madhi, Adrian V S Hill, Andrew M Prentice, Philip Bejon, Gibran Hemani, George Davey Smith, Manjinder S Sandhu, Alison M Elliott, Thomas N Williams, Adebowale Adeyemo, Sarah H Atkinson

## Abstract

Iron is an essential trace element for both humans and pathogens, but its genetic determinants are understudied in Africa where iron deficiency and infectious diseases are highly prevalent. We conducted genome-wide association studies for six iron-related biomarkers in 3928 children from five sites across Africa with replication in 2868 African American adults. We identified previously unreported loci for transferrin in *GTF3C5,* a gene regulating cellular iron-uptake; for soluble transferrin receptor in *FREM3*, the Dantu locus; and for hepcidin in *CHCHD7*/*SDR16C5*. The lead *GTF3C5* (rs2905094) and *FREM3* (rs141274959) variants were both associated with protection against severe malaria and against bacteremia in large case-control studies. The *CHCHD7*/*SDR16C5* lead variant, rs73596248, increased hepcidin concentrations and protected against bacteremia. We report limited transferability of polygenic risk scores derived from European ancestry studies to African populations. Our findings advance the understanding of the genetics of iron status in Africa and suggest an important link between iron and infection.

## Introduction

Iron is an essential nutrient that is required for critical biological functions in almost all living organisms. Iron deficiency affects over half of young African children,^1^ and is a leading cause of years lived with disability.^2^ Iron status influences susceptibility to infection since pathogens require iron to proliferate, but iron is also essential for host immunity.^3,4^ This is important in the context of Africa due to the high infectious disease burden. In 2023, there were an estimated 263 million cases of malaria, and 597,000 deaths globally with 95% of these occurring in the WHO African region.^5^ Similarly, age standardised mortality rates due to invasive bacterial infections were highest in sub-Saharan Africa with an estimated 230 deaths per 100 000 population in 2019.^6^

Iron plays a crucial role in the relationship between host and pathogen,^7^ and the high infectious disease burden in Africa might have created different evolutionary pressures on iron-regulating genes. Iron homeostasis is tightly regulated in humans since both iron deficiency and overload increase the risk of infection.^8^ Iron is tightly sequestered by transferrin and lactoferrin to reduce its availability to systemic organisms, while cellular iron uptake through transferrin receptor 1 is critical for the development of host immunity.^9^ During inflammation and infection iron availability is further reduced by the actions of hepcidin which inhibits enteric absorption and sequesters plasma iron into macrophages^7,10^ and the liver thus inhibiting malaria parasites and bacteria.^11,12^ Iron is also critical to host adaptive immunity.^13^

The genetic architecture of iron status in African populations remains largely understudied, in contrast to the many large-scale genome-wide association studies (GWAS) of iron status conducted in European ancestry populations.^14,15^ Those studies that are available in Africa are largely limited to candidate variant studies^16,17^ and to our knowledge, no iron GWAS has been undertaken in continental African populations. A GWAS of iron status was conducted in 2347 middle-aged African-American adults,^18^ however transferability to continental African populations might be complicated by admixture, by limited representation of the greater genetic diversity found among continental Africans,^19,20^ and by the profoundly different environmental exposures in terms of nutrition and infectious disease.^21,22^ Even within Africa, there exist divergent patterns in LD which might yield region-specific patterns of association.^23^ Given the greater genetic variation in Africans,^19^ GWAS may have better resolution in African samples, and because of distinct patterns of infection, an iron GWAS may reveal differential effects compared to other populations due to genotype-environment interactions.

To enhance discovery of genetic determinants of iron status in an understudied population, we conducted a GWAS of six iron biomarkers (transferrin, hepcidin, soluble transferrin receptor (sTfR), ferritin, serum iron, and transferrin saturation (TSAT)) in 3,928 children from Kenya, Uganda, Burkina Faso, The Gambia and South Africa with replication in 2,868 African American adults. We identify three previously unreported African-specific genome-wide significant loci that are associated with iron homeostasis and risk of severe malaria and invasive bacterial infections.

## Results

### Characteristics of discovery sample

To assess the genetic contribution to iron status in continental Africans, we performed a discovery GWAS analysis using data from five sites in Africa (Fig. 1a).^24^ A total of 3928 children (1059 Kenyan, 1360 Ugandan, 348 Burkinabe, 611 South African, and 550 Gambian), aged between birth and eight years were included. The characteristics of the 3928 children are summarized in Supplementary Table 1. Prevalence of iron deficiency, and geometric mean concentrations of measured iron biomarkers (transferrin, ferritin, sTfR, hepcidin, serum iron, and TSAT) varied by study site (Fig. 1b). Inflammation, malaria parasitemia and undernutrition were highly prevalent among study participants (Supplementary Table 1). Principal component plots of the genotypes showed clustering of the study participants by geography and ethnolinguistic group (Fig. 1c).

**Fig 1.**
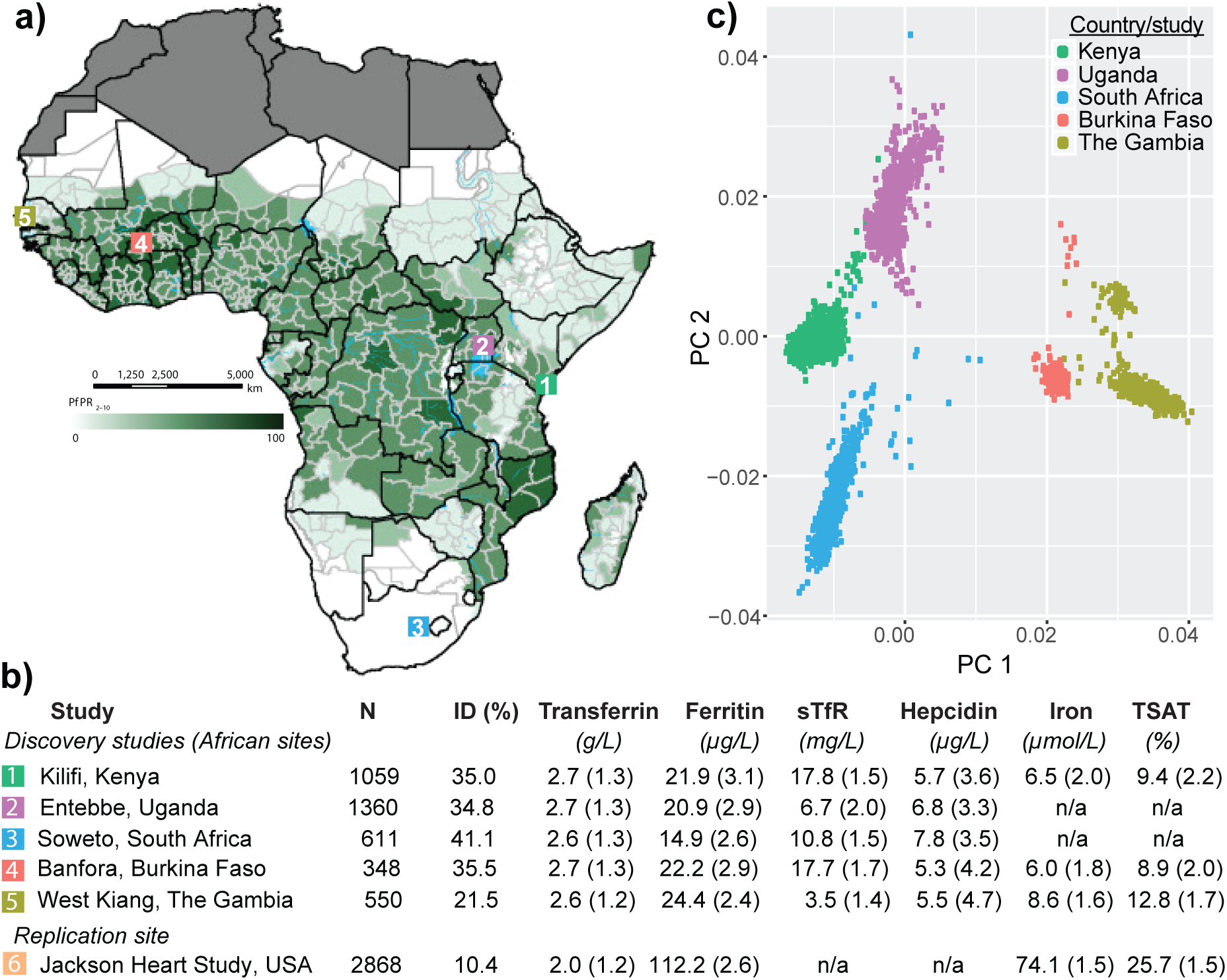
Characteristics of study populations. **a)** Africa malaria map showing the location of the discovery sites included in the current study and predicted posterior predictions of age-standardized *P. falciparum* prevalence (PfPR_2–10_) taken from Snow *et al* 2017.^50^ **b)** Prevalence of iron deficiency (ID) and geometric means of iron markers by study site. Iron deficiency was defined as serum ferritin <12 µg/L with no inflammation, or <30 µg/L in the presence of inflammation (C-reactive protein>5mg/L or α1-antichymotrypsin >0.6g/dL) in children <5 years or <15µg/L with no inflammation or <70 µg/L in the presence of inflammation in children ≥5 years).^51^ **c)** Principal components plot of genotypes of the discovery study participants. sTfR, soluble transferrin receptor; TSAT, transferrin saturation; PC (Principal Components); n/a, not available.

A summary of quality control steps of genotypes for each population is shown in Supplementary Table 2. The number of variants imputed into individual datasets is shown in Supplementary Table 3. Only high-quality SNPs (imputation information score >0.3) with a minor allele frequency (MAF) >0.01 were included in genetic association analyses.

### Discovery genetic association analyses

To identify variants associated with iron biomarkers, we performed individual GWAS within each of the African discovery sites (Supplementary Fig. 1) followed by meta-analysis. Fig. 2 shows the distribution of association statistics for the GWAS of the six iron biomarkers after fixed-effects meta-analyses of all five African sites. We applied a linear mixed model including a genetic relatedness matrix (GRM) to account for relatedness in the sample and age and sex as covariates and found minimal population stratification or inflation of the association statistics (lambda ranged from 0.98 to 1.05, Supplementary Fig. 2). Additional adjustment for inflammatory markers yielded similar results. Meta-analysis of GWAS results from all the African cohorts yielded a previously reported genome-wide significant locus for transferrin on chromosome 3 at the *TF* gene (Fig. 2a) and a previously unreported African-specific locus associated with hepcidin levels (Fig. 2b). We did not observe any loci at genome-wide significance (set at P<5×10^-8^) after meta-analysis of all GWAS results for ferritin, sTfR, serum iron, and TSAT from all African study sites (Figs. 2c, 2d, 2e and 2f).

**Fig 2.**
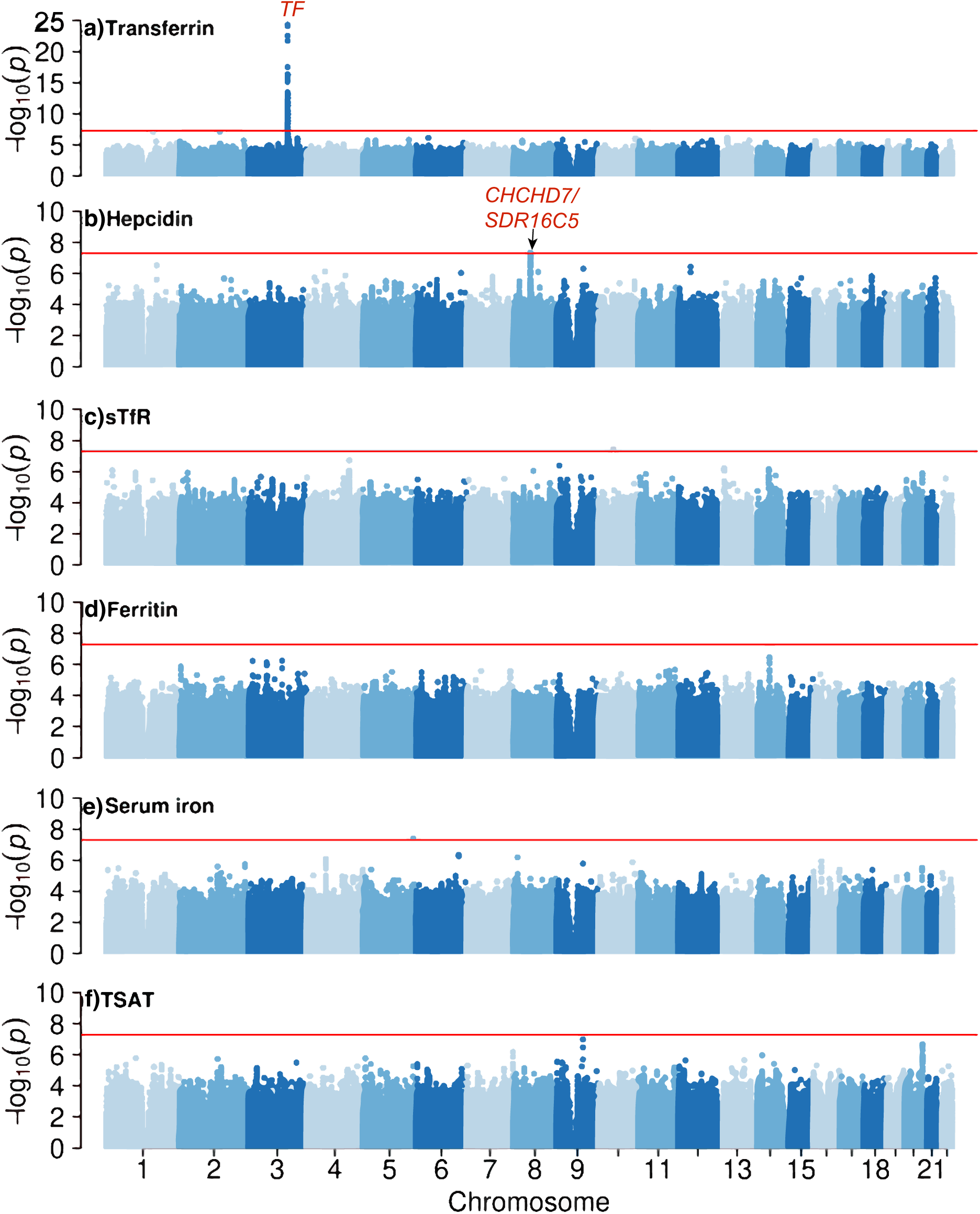
Manhattan plots of meta-analysis results of discovery samples. Meta-analysis included all five African sites except for serum iron and TSAT which were missing in Ugandan and South African samples. Genome-wide significant signals are annotated *TF*, for transferrin and *CHCHD7*/*SDR16C5* for hepcidin. Genetic variants were tested for association with iron biomarkers per study site using linear mixed models and evidence was combined across sites using inverse-variance weighted meta-analysis in a fixed-effects framework. The red line indicates threshold for genome-wide significance (set at 5×10^-8^).

The *TF* locus is well characterized across ancestries. The lead SNP in our African transferrin GWAS was rs6762719 (P=1.59×10^-27^) which is in strong LD (r^2^=0.9) with the lead common SNP (rs3811647) associated with transferrin levels in European ancestry transferrin GWAS.^25^ The rs3811647 SNP (P= 3.45×10^-16^) was among the top SNPs in our African transferrin GWAS. After conditional analyses adjusting for rs6762719, we found another independent locus (lead SNP rs4854748) indicating at least two independent loci for transferrin (Table 1) as previously reported in African-American participants.^18^ This SNP (rs4854748) was in strong LD (r^2^=0.9) with the lead SNP, rs9872999, in the conditional analysis in African-Americans.^18^ Notably, rs4854748 has been strongly associated with reduced total iron binding capacity (TIBC), a proxy for transferrin levels, in the largest European-ancestry TIBC GWAS (P=3.05×10^-578^).^15^ Genome-wide significant SNPs associated with transferrin levels in our discovery meta-analysis replicated in the Jackson Heart Study (JHS) and the largest European-ancestry TIBC GWAS^15^ (except rs113656013 which is rare in Europeans (minor allele frequency = 0.001 compared to 0.17 in Africans); Supplementary Table 4).

**Table 1.** Top independent SNPs associated with iron biomarkers in African children.

| SNP | Chr | Hg19 BP | EA | OA | Gene | EA Freq | Beta | Se | P |
| --- | --- | --- | --- | --- | --- | --- | --- | --- | --- |
| *Transferrin (all African sites)\** |  |  |  |  |  |  |  |  |  |
| rs6762719 | 3 | 133480817 | G | A | *TF* | 0.25 | 0.28 | 0.03 | 1.59x10<sup>-27</sup> |
| rs4854748 | 3 | 133446881 | T | C | *TF* | 0.20 | -0.24 | 0.03 | 1.12x10<sup>-16</sup> |
| *Transferrin (Uganda and Kenya)* |  |  |  |  |  |  |  |  |  |
| rs2905094 | 9 | 135902689 | T | C | *GTF3C5* | 0.16 | -0.23 | 0.04 | 8.08x10<sup>-9</sup> |
| *Hepcidin (all African sites)* |  |  |  |  |  |  |  |  |  |
| rs73596248 | 8 | 57134187 | A | G | *CHCHD7/*<br>*SDR16C5* | 0.06 | 0.27 | 0.05 | 4.51x10<sup>-8</sup> |
| *sTfR (Kenya)* |  |  |  |  |  |  |  |  |  |
| rs552439837 | 4 | 144446525 | G | A | *FREM3* | 0.09 | 0.48 | 0.09 | 2.78x10<sup>-8</sup> |
| Chr, chromosome; Hg19 BP, human genome build 19 base pair; EA, effect allele; OA, other allele; EA Freq, effect allele frequency; Se, standard error; P, two-sided P value from an additive model.<br>*R<sup>2</sup> between rs6762719 and rs4854748 was 0.03 in the African populations. |  |  |  |  |  |  |  |  |  |

### *GTF3C5*, an East African-specific locus for transferrin

Since genetic associations can be highly population-specific across Africa, we performed meta-analysis by African region (i.e., East, West and South Africa; Supplementary Fig. 3). For transferrin levels, we observed a previously unreported East African (Kenya and Uganda) specific locus on chromosome 9 (Fig. 3a). Three SNPs reached genome-wide significance in East Africa only (Supplementary Table 5). The lead SNP, rs2905094, was in high LD with the other two SNPs (Fig. 3b) and explained 1.42% of variation in transferrin levels. The T allele frequency of rs2905094 in East Africa was 16% with allele frequencies of 14-23% across the African sites and in AfricanAmericans in the JHS, and of 33% in Europeans (Fig. 3c). The rs2905094 SNP was also associated with increased levels of ferritin, TSAT and serum iron, decreased levels of sTfR (Fig. 3d), and a 19% reduction in iron deficiency (OR=0.81, (95% CI 0.68, 0.97); P=0.02) in East Africa.

**Fig 3.**
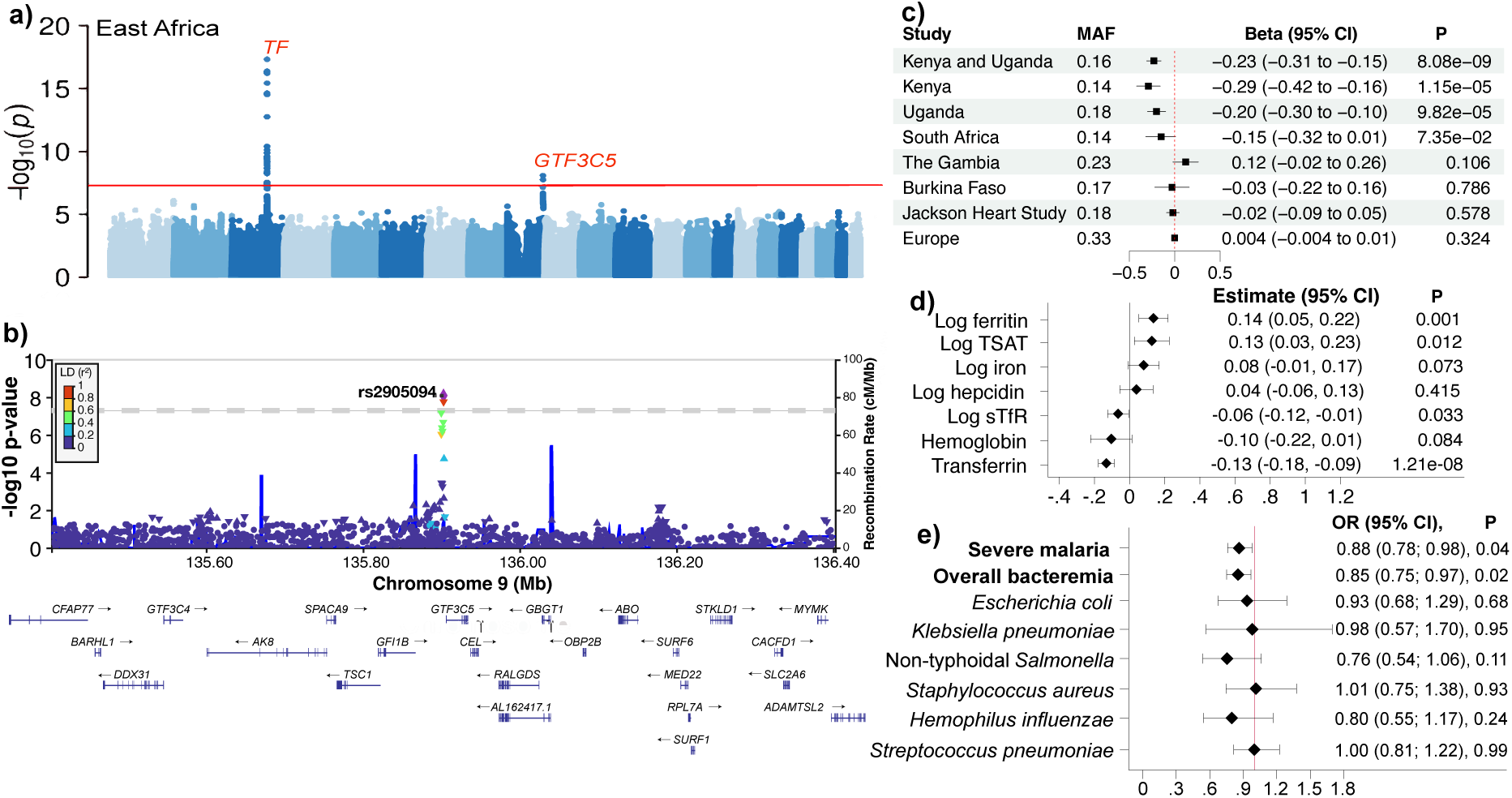
*GTF3C5* is a previously unreported locus associated with transferrin levels, severe malaria and bacteremia in East Africa. **a)** Meta-analysis of transferrin GWAS results in East Africa (Kenya and Uganda). **b)** A regional plot of the *GTF3C5* locus in East Africa. The most strongly associated variant, rs2905094 (P=8.08×10^-9^), is represented by a purple diamond. Additional SNPs are colored by their correlation (r^2^) with rs2905094. Location and direction of transcription of protein-coding genes within the region are shown. **c)** A forest plot by study site showing the minor allele frequency (MAF), beta and 95% confidence interval, and additive model two-tailed GWAS p-values of the association between rs2905094 and transferrin levels. **d)** Association between rs2905094 and other iron biomarkers in Kenyan and Ugandan children. Linear regression models were adjusted for age, sex, and study site. Estimates were derived from an additive model. **e)** A forest plot showing odds ratios of rs2905094 for risk of severe malaria, overall bacteremia, and common bacterial isolates in East African children.

We then used external reference data to annotate and evaluate the functional or regulatory impact of genome-wide significant variants and nearby gene(s). The three genome-wide significant SNPs (Supplementary Table 5) mapped to the *GTF3C5* (general transcription factor IIIC, polypeptide 5) gene and were all intronic. Using the African Functional Genomics Resources (AFGR, https://github.com/smontgomlab/AFGR), rs2905094 was a significant eQTL for *GTF3C5* (Beta = 0.07, se =0.03, P=0.04, n=593) in immortalized lymphoblastoid cell line. *GTF3C5* is involved in RNA polymerase III-mediated transcription. It is an integral and tightly associated component of the DNA-binding transcription factor IIIC subcomplex that directly binds tRNA and virus-associated RNA promoters. *GTF3C5* is expressed ubiquitously, with a relatively higher expression in the cerebellum. *GTF3C5* knockdown has been shown to increase endocytosis and internalization of transferrin.^26^

Given the above observations, we next performed haplotype analyses to investigate why the *GTF3C5* locus association was East African-specific. We found two dominant haplotypes from chromosomes with the rs2905094 mutant (T) allele (Supplementary Fig. 4). Haplotype 1 accounted for approximately 70% and 60% of chromosomes in The Gambia and Burkina Faso, respectively and less than 20% in South Africa, Uganda and Kenya. Haplotype 2 accounted for 40%, 25% and 15% of chromosomes from Kenya, Uganda, and South Africa, respectively, but was absent in The Gambia and Burkina Faso. Given these population differences, we hypothesized that haplotype 2 might be responsible for the East-African specific transferrin signal. Consistent with this hypothesis, we found that rs2905094 had the largest effect size in Kenya, followed by Uganda, but was not associated with transferrin levels in South Africa, Burkina Faso, and The Gambia (Fig. 3c). This suggested that rs2905094 might be in LD with an unidentified causal SNP within the East-African specific haplotype 2.

Since transferrin levels influence risk of malaria and bacterial infections,^27,28^ we investigated the association between rs2905094 and these infections using large case-control GWAS of severe malaria (7957 cases and 7746 controls)^29^ and bacteremia (1536 cases and 2677 controls).^30,31^ We compared fixed effects meta-analyses of severe malaria GWAS data from East Africa (i.e. Kenya, Malawi, and Tanzania) and West Africa (The Gambia, Mali, Burkina Faso, Ghana, Nigeria, and Cameroon; Supplementary Fig. 5) since the effect of rs2905094 on transferrin levels was restricted to East Africa. We found that rs2905094 was associated with 12% (OR=0.88, (95% CI 0.78, 0.98) protection against severe malaria in East Africa where it had an impact on transferrin levels but was not associated with protection in West Africa where it had no impact on transferrin levels (OR=0.95; 95% CI 0.87, 1.03 (Fig. 3f and Supplementary Fig. 5). The rs2905094 was associated with 15% (OR=0.85, (95% CI 0.75, 0.97; P=0.02) protection against overall bacteremia in Kenyan children (Fig. 3f).

### *CHCHD7/SDR16C5*, an African-specific locus for hepcidin

Meta-analysis of hepcidin GWAS results from all five African sites yielded a previously unreported locus at genome-wide significance, found on chromosome 8 near the *CHCHD7* (Coiled-Coil-Helix-Coiled-Coil-Helix Domain Containing 7) and *SDR16C5* (Short Chain Dehydrogenase/Reductase Family 16C Member 5) genes (Fig. 2b). The regional plot suggests that this association is within an extended haplotype (Fig. 4a). This region shows evidence of positive selection among two 1000G African populations (Luhyas (Kenya) and Yorubas (Nigeria)); iHS = 1.5, XP-EHH =2.5, Fay & Wu’s H =-7.6, and Tajima’s D =-2.8) (https://pophumanscan.uab.cat/tables.php). The A allele of the lead SNP, rs73596248 occurred at a frequency of 6% in Africans with consistently increased hepcidin levels across all the African sites (Fig. 4b). rs73596248 explained 0.82% of variation in hepcidin levels. In contrast, the rs73596248 A allele was very rare in European and Latin American populations and was monomorphic in Asian populations (Supplementary Table 6). rs73596248 was associated with a 21% (OR=0.79, (95% CI 0.64, 0.97); P=0.03) protection against iron deficiency, but was not significantly associated with individual iron biomarkers (Supplementary Fig. 6a).

**Fig 4.**
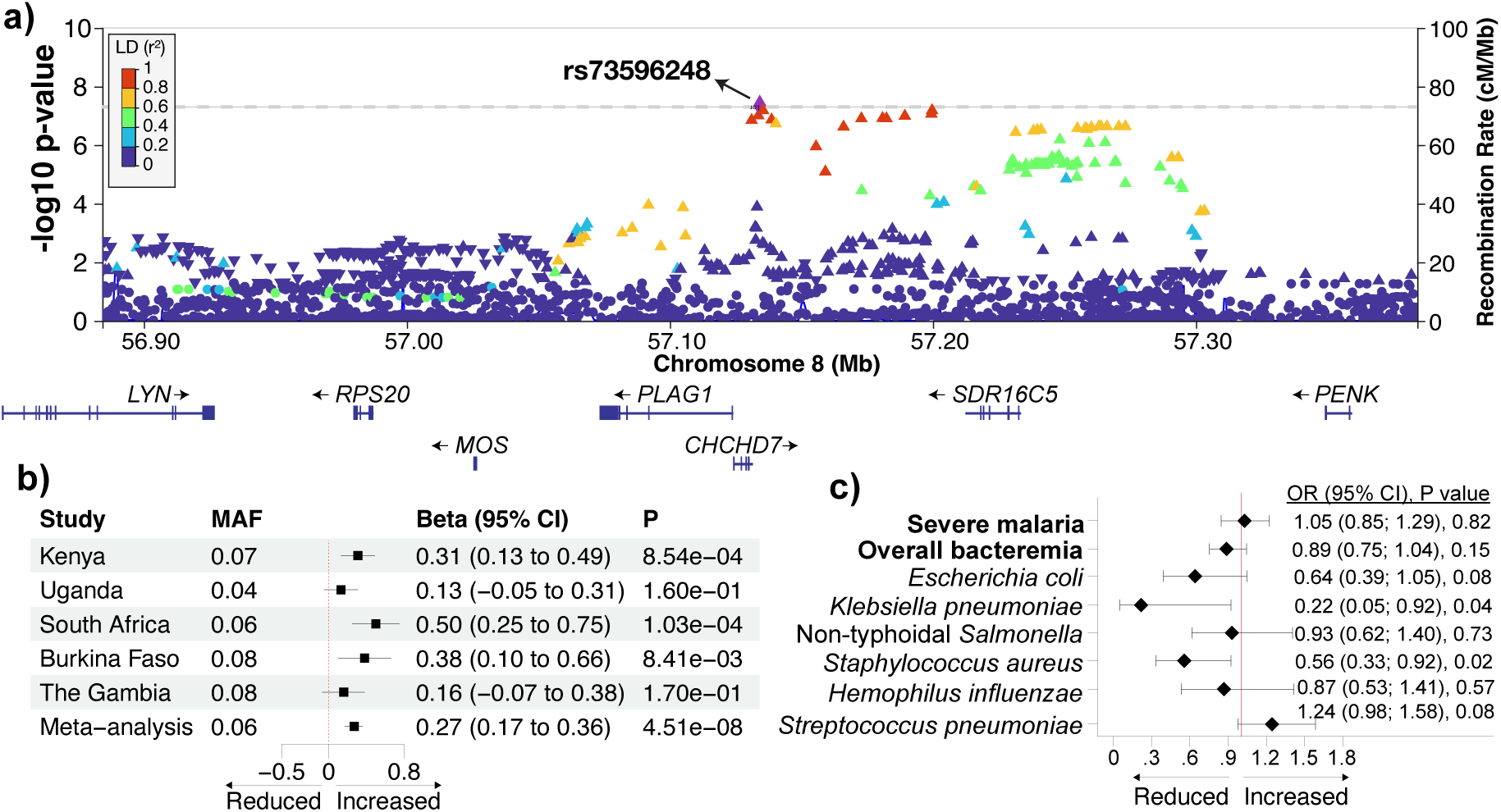
A previously unreported locus associated with hepcidin levels in African populations. **a)** A regional plot for the locus. The most strongly associated variant, rs73596248 (P=4.51×10^-8^), is represented by a purple diamond. Additional SNPs are colored by their correlation (r^2^) with rs73596248. Location and direction of transcription of protein-coding genes within the region are shown. **b)** A forest plot by study site showing the minor allele frequency (MAF), beta and 95% confidence interval, and additive model two-tailed GWAS p-values of the association between rs73596248 and hepcidin levels. Hepcidin data was not available in the Jackson Heart Study so replication in that population was not tested. **a)** A forest plot showing the odds ratios of rs73596248 and severe malaria, overall bacteremia, and common bacterial isolates.

Based on available and limited evidence, rs73596248 is downstream of *CHCHD7* and is an intergenic variant between the *CHCHD7* and *SDR16C5* genes. rs73596248 is a significant eQTL for *CHCHD7* (P=1.50×10^-4^) in the AFGR. *CHCHD7* is a protein coding gene predicted to be in the mitochondrial intermembrane space and is overexpressed in fibroblasts. The *SDR16C5* gene encodes short chain dehydrogenase reductase which oxidizes retinol to retinoic acid and is overexpressed in the lung. Retinoic acid has been reported to suppress production of hepcidin.^32^ Although the *CHCHD7/SDR16C5* locus has been strongly associated with height, we did not find an association between rs73596248 and anthropometric indices including height in our African cohorts (Supplementary Fig. 7).

Since hepcidin is implicated in the pathophysiology of malaria and bacterial infections,^11,12,33^ we determined the association between rs73596248 and these infections in the largest case-control GWAS of severe malaria and bacteremia mentioned above. We found that rs73596248 was associated with protection against bacteremia due to *Klebsiella pneumoniae* (OR=0.22; 95% CI 0.05, 0.92; P=0.04) and *Staphylococcus aureus* (OR=0.56; 95% CI 0.33, 0.92; P=0.02). Protection against overall bacteremia and severe malaria did not reach statistical significance (Fig. 4c).

### *FREM3*, a Kenyan-specific locus for soluble transferrin receptor

We observed a genome-wide significant locus at the *FREM3* (FRAS1 Related Extracellular Matrix 3) gene that was associated with an increase in sTfR levels in Kenya (Fig. 5a). *FREM3* is within the complex Dantu region that is under strong selective pressure for protection against malaria along the Kenyan coast.^34,35^ The regional plot suggests an extended haplotype underlying the association (Fig. 5b). The top SNPs were rare or monomorphic outside Kenya (Supplementary Table 7). The G allele of the lead SNP, rs552439837, occurred at a frequency of 9% in Kenya (Table 1) and was also associated with an increase in TSAT and serum iron concentrations (Supplementary Fig. 6b). rs552439837 explained 3.77% of variation in sTfR levels. The causal malaria-related Dantu SNP, rs186873296, ^35^ was in strong LD (r^2^>0.9) with rs552439837 and was also associated with increased sTfR levels (effect size=0.34, se=0.07, P= 1.3×10^-6^, G allele frequency=11.3%). Owing to the Kenyan-specific nature of the Dantu region, we did not find any reported eQTL data for the top sTfR GWAS SNPs in GTex or AFGR databases.

**Fig 5.**
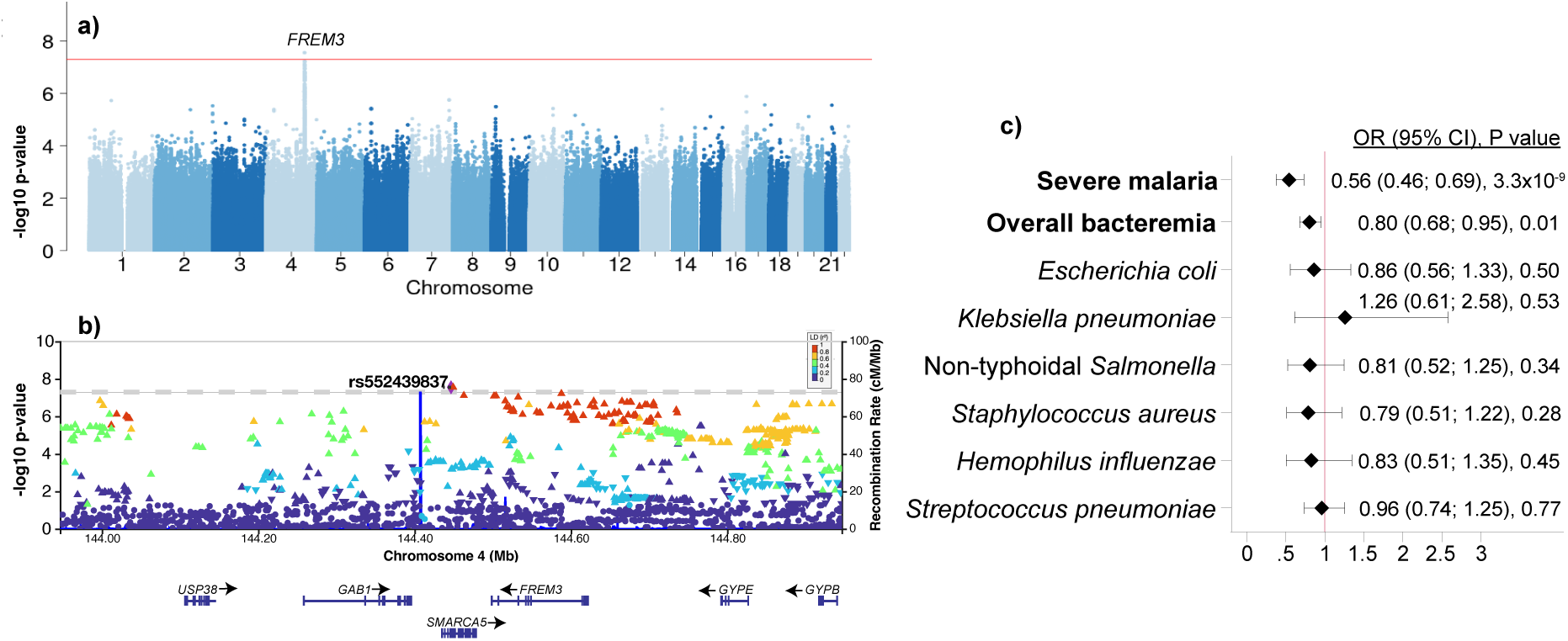
Genome-wide association study of soluble transferrin receptor in Kenya. **a)** Manhattan plot of GWAS of soluble transferrin receptor (sTfR) in Kenya. **b)** A regional plot showing correlation between the lead SNP, rs552439837 (purple diamond) and SNPs within the region. **c)** A forest plot showing odds ratio of rs141274959 and severe malaria, overall bacteremia, and common bacterial isolates. rs141274959 had an r^2^=0.9 with rs552439837 which was missing in the malaria and bacteremia GWAS.

The lead SNP, rs552439837 was missing in the large case-control GWAS of severe malaria and bacteremia. We therefore identified the closest available SNP, rs141274959, from among the genome-wide significant sTfR GWAS SNPs (Supplementary Table 7). rs141274959 is in strong LD with rs552439837 (r^2^=0.9) and was highly protective against severe malaria (OR=0.56; 95% CI 0.46, 0.69; P=3.3×10^-9^) and bacteremia (OR=0.80; 95% CI 0.68, 0.95; P=0.01; Fig. 5c).

### Transferability of established European iron GWAS SNPs

To investigate the transferability of European iron GWAS findings to Africans, we identified the estimates of index SNPs that were associated with iron biomarkers in the largest GWAS of European-ancestry populations^14,15^ in our meta-analyzed GWAS results of African children (Supplementary Table 8 and 9). We considered a GWAS result to be replicated if the estimate in the African iron GWAS was in the same direction as the estimate in the European iron GWAS, and if the association in African iron GWAS was statistically significant (P< 0.05). Most of the variant allele frequencies differed substantially between European ancestry and continental African populations (Supplementary Table 8 and 9). The *TF* locus replicated while the other loci had limited replication. Notably, the *HFE* variants (rs1800562 and rs1799945) could not be replicated since the associated SNPs were very rare or monomorphic in Africans. The *TMPRSS6* variant, rs855791, replicated for TSAT and had similar effect size for the other iron biomarkers in Africans and Europeans (Supplementary Table 8 and 9). However, the effect allele (A) frequency of rs855791 is lower in African (7.7%) compared to European (43.1%) populations. In summary, of the 138 index SNPs available in our African iron GWAS, 69 (25 for ferritin, 20 transferrin, 9 serum iron, 6 hepcidin, 6 sTfR, and 4 TSAT) had the same direction of effect as in the estimate in the European iron GWAS but only the *TF* locus had a statistically significant P value (Supplementary Table 8 and 9). We then calculated polygenic risk scores (PRS) for each iron biomarker using the European iron biomarkers GWAS index SNPs and similarly found that only the transferrin PRS replicated in the current study (Fig. 6).

**Fig 6.**
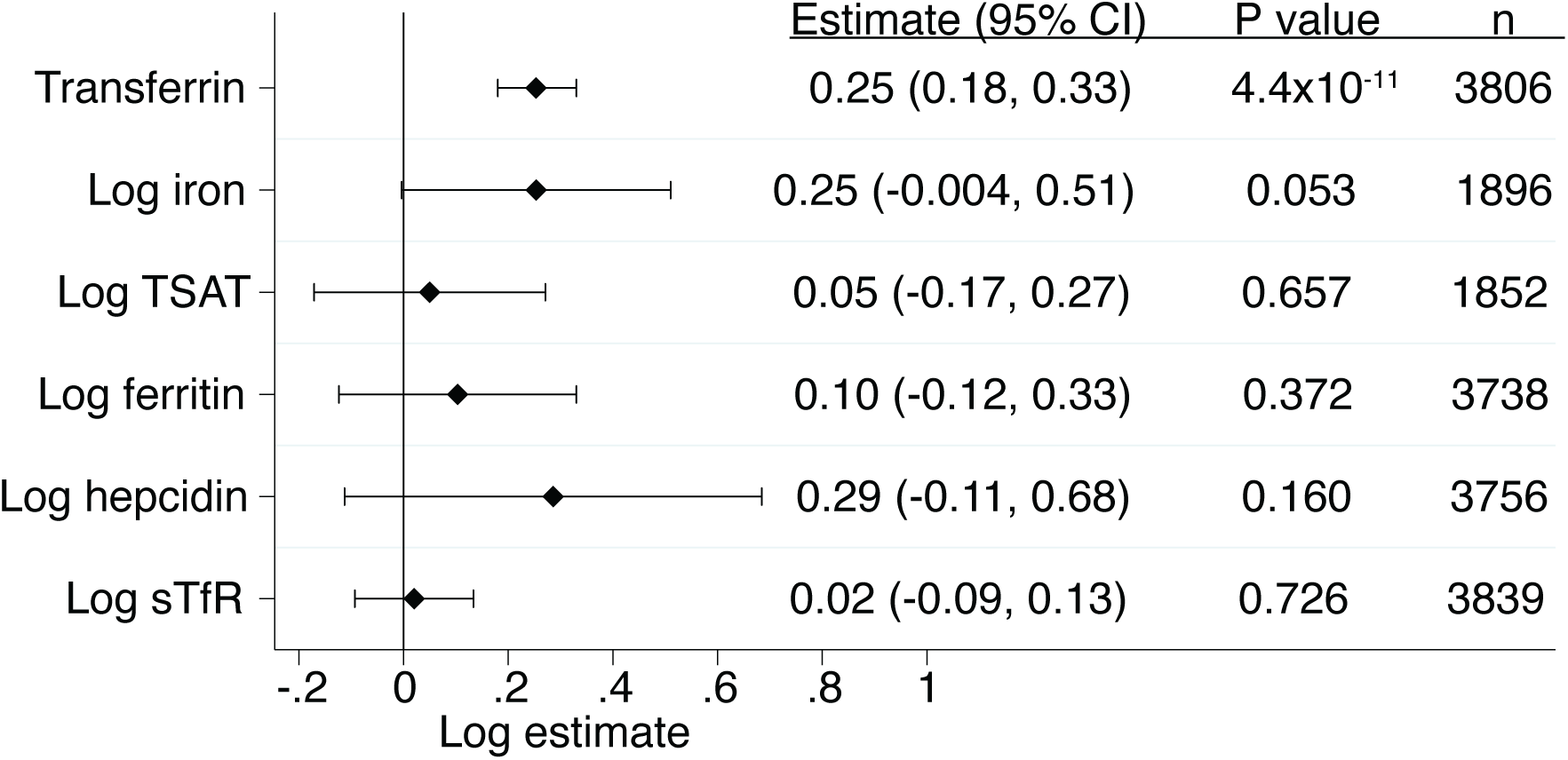
Transferability of polygenic risk scores (PRS) derived from European ancestry iron GWAS lead SNPs to continental African populations. Association between European ancestry PRS and iron biomarkers in African children. Regression models were adjusted for age, sex, and study site. For each iron biomarker, we calculated a PRS weighted using effect sizes reported in European ancestry iron biomarker GWAS and reflecting an increase in levels of the biomarker (included SNPs are shown in Supplementary Table 8 and 9). In our African populations, we used transferrin as a proxy measure for total iron binding capacity (TIBC), which was reported in the European GWAS. TIBC indicates the amount of iron that can be bound to transferrin. TSAT=transferrin saturation; sTfR, soluble transferrin receptor.

## Discussion

African populations are substantially under-represented in human genomic studies.^36,37^ In this study, we report the first GWAS of iron status in continental African populations. We replicated a previously reported signal at the *TF* gene that was associated with transferrin levels.^18,25^ We further discovered three African-specific genetic loci that influence iron status in African children. We identified a previously unreported East African specific locus, *GTF3C5,* implicated in intracellular iron uptake, which was associated with reduced transferrin levels and protection against malaria and bacteremia. We also found a previously unreported locus within the *CHCHD7/SDR16C5* region, associated with increased hepcidin levels and protection against bacteremia. The malaria protective Dantu locus,^35^ was associated with increased sTfR levels in Kenyan children and protection against bacteremia. These region-specific findings underscore the existence of divergent patterns in LD among African populations.^23^ We further found limited transferability of findings from European iron GWAS to African populations. Africa’s diverse lifestyle, cultural and environmental factors such as long-term burden of infections might have shaped the genomic architecture of its people.

Transferrin, synthesised in the liver, is the major iron-binding and transport protein that delivers iron to cells. It is highly conserved in humans and plays an important role in infection by sequestering iron away from pathogens, contributing to ‘nutritional immunity’.^38^ Our transferrin GWAS analyses yielded three independent loci: two at the *TF* gene and one novel locus at the *GTF3C5* gene. The two independent loci at the *TF* gene have previously been reported and are well characterised across ancestries.^15,25,39,40^ The *GTF3C5* locus was associated with reduced transferrin levels and is specific to East African populations. *GTF3C5* encodes transcription factor IIIC63 (TFIIIC63) which binds to DNA to recruit transcription factor IIIB and RNA polymerase III to mediate the transcription of small noncoding RNAs, such as tRNAs. *GTF3C5* has been implicated in transferrin endocytosis.^26^ Through a phenotypic profiling of genes involved in transferrin endocytosis by genome-wide RNAi screening using a quantitative multiparametric image analysis, *GTF3C5* gene knockdown caused increased endocytosis and internalization of transferrin.^26^ We observed SNPs at *GTF3C5* that were associated with a decrease in plasma transferrin levels suggesting that the SNPs might cause an increase in endocytosis and internalization of transferrin similar to the *GTF3C5* gene knockdown. We also found that the lead SNP, rs2905094 was associated with an increase in levels of ferritin, serum iron, and TSAT and a decrease in sTfR levels, in keeping with improved iron status. Indeed, we found that rs2905094 was associated with protection against iron deficiency suggesting that the variant might play a role in iron absorption.

The *GTF3C5* locus appears to influence iron status in East Africa only despite the effect allele occurring at a similar frequency in other African ancestry populations and an even higher frequency (33%) in Europeans. This finding might be explained by multiple factors. Owing to the distinct demographic histories of populations from different geographic regions in Africa, this locus might be characterized by distinct haplotype block structures. We found that East African populations displayed a unique haplotype suggesting that an unknown causal SNP might be in LD with our index SNPs in East African populations but not in other populations. Gene environment interactions could also be at play since we found that the locus was associated with severe malaria in East Africa but was not associated with protection in West Africa. This population-specific finding further underscores the high diversity of African genomes and existence of divergent patterns in LD among African populations.^19,23^

The *GTF3C5* locus was further associated with protection against severe malaria and bacteremia suggesting a link between iron metabolism and infection. We hypothesize three possible mechanisms. Firstly, an increase in intracellular iron within immune cells or improved iron status might play a role in improved immunity thereby protecting against infection.^4,41^ Using a transgenic mouse model carrying a mutation in the transferrin receptor (thus limiting the ability of cells to internalise iron), Wideman et al reported impaired immune response and increased malaria parasitemia.^41^ It is therefore possible that increased internalised iron improves immunity against pathogens. Secondly, by increasing endocytosis of transferrin-bound iron,^26^ excess of iron within cells is potentially toxic to intracellular pathogens.^42^ Indeed, we found that the variant was associated with protection against severe *P. falciparum* an intracellular pathogen. Thirdly, since the malaria parasite can acquire iron bound to transferrin,^28^ a reduction in transferrin might deny the parasite of iron for growth and multiplication. Further research is required to test these hypotheses.

We found a previously unreported African specific locus between the *CHCHD7* and *SDR16C5* genes that was associated with increased hepcidin levels in populations across Africa. SNPs within this locus occurred at a frequency of ∼6% in Africans but were very rare or monomorphic in non-African populations. Concordantly, we found evidence that the locus is under positive selective pressure probably to protect against pathogens. Hepcidin is the master regulator of systemic iron homeostasis and plays a crucial role in the body’s response to infection by inducing ‘nutritional immunity’.^10,33^ The lead SNP, rs73596248, was associated with increased hepcidin levels and protection against *K. pneumoniae* and *S. aureus* bacteremia. This finding is also in agreement with our previous report that severely anemic children are unable to reduce hepcidin in response to infection and have increased risk of bacteremia.^43^

We found that the malaria-protective Dantu region at the *FREM3* locus was associated with increased sTfR levels and protection against bacteremia in Kenyan children. This locus is unique to the Kenyan population since SNPs were rare or monomorphic elsewhere. *FREM3* is in linkage disequilibrium with a complex structural mutation within the glycophorin gene region (comprising *GYPA*, *GYPB*, and *GYPE*) that encodes for the rare Dantu blood group antigen and is associated with protection against severe malaria in East Africa.^35^ In our analyses, the locus was associated with protection against both severe malaria and bacteremia. The protection against bacteremia is not surprising since malaria infection strongly predisposes individuals to bacteremia in malaria-endemic areas.^44^ Recent analyses from the same population showed that the Dantu polymorphism, rs186873296, is protective against bacteremia via its effect on malaria risk.^45^ sTfR is a marker of erythropoiesis or younger RBCs and when increased it indicates iron deficiency. In an observational study, we previously showed that iron deficiency was protective against malaria infection.^46^ This finding might also suggest that Dantu RBCs are younger than non-Dantu RBCs as previously shown.^47^ However, further research is required to elucidate the complex interplay between iron and malaria infection.

We further found limited transferability of European iron GWAS findings to continental African populations. Using index variants identified in the largest iron GWAS in European populations,^14,15^ we were only able to formally replicate other prior reported loci in the *TF* locus, in African children. Notably, the *HFE* polymorphisms were very rare or monomorphic in Africans and it is well known that the hemochromatosis-causing variants are not observed in African populations. The *TMPRSS6* SNP, rs855791, although not as common in African compared to European populations (allele frequency of 7.7% in Africans versus 43.1% in Europeans), replicated in terms of effect size but not significance (P>0.05) except for TSAT. Considering the low frequency of rs855791 in Africans, it is likely that we were underpowered to observe a statistically significant association. The *HFE* and *TMPRSS6* polymorphisms are often used in Mendelian randomization (MR) studies involving European populations,^48,49^ but would have limited application in African MR and functional studies. All the other loci reported in European iron GWAS did not replicate (in terms of both direction of effect and P values) in our study populations. Similarly, only the transferrin PRS replicated in our African iron GWAS. However, our sample size was small compared to the large sample sizes reported in European GWAS. The European GWAS were also conducted in adults while the current study involved children, and regulation of iron homeostasis could differ across the life course. Moreover, most variants had very different effect allele frequencies between African and European populations which might further explain the limited transferability and a need for African genomic studies.

The strengths of the present study include a focus on diverse understudied continental African ancestral groups and inclusion of a wide range of iron biomarkers. To our knowledge, this study is the first GWAS of iron biomarkers within continental Africa. We have identified previously unreported African-specific loci and further determined their impact of on infections using the largest case-control studies of severe malaria and bacteremia. A potential limitation is that variants with small effect sizes were not detectable with the present sample size. To overcome this limitation, there is a need for larger genomic studies of iron status within continental Africa.

In conclusion, we found African-specific genetic loci that are associated with iron status in African children. Malaria and bacteremia may have shaped the genetic architecture of iron status in Africans. However, mechanistic studies using the identified loci are required to infer the causal relationship between iron and pathogens. Finally, we found limited transferability of European iron GWAS to Africans. These findings further underscore the importance of conducting genomic studies in Africans to better address specific medical needs and global health inequalities.

## Data Availability

All data are available in the main text or the Supplementary Materials. All direct genotypes from individuals after QC along-side imputed data have been submitted to the European Genome-Phenome Archive under accession, with the datasets under EGAD00010002578 and EGAD00010002583 (Uganda); EGAD00010002582 and EGAD00010002580 (South Africa); EGAD00010002581 and EGAD00010002579 (Burkina Faso). Applications for access to primary individual level de-identified phenotype data for the Kilifi, Kenya; Entebbe, Uganda; Banfora, Burkina Faso; and West Kiang, The Gambia cohorts can be made through the Data Governance Committee.

## Methods

### Ethical Approvals

Individual study site ethical approvals were obtained. For Kilifi, Kenya (by the Scientific Ethics Review Unit of the Kenya Medical Research Institute (KEMRI/SERU/CGMR-C/046/3257/2983)); Entebbe, Uganda (locally by the Uganda Virus Research Institute (GC/127/12/07/32) and Uganda National Council for Science and Technology (MV625), and in the UK by the London School of Hygiene and Tropical Medicine (A340) and Oxford Tropical Research (OTR) (39-12, 42-14 and 37-15) Ethics Committees); Banfora, Burkina Faso (by Ministere de la Recherche Scientifique et de l’Innovation in Burkina Faso (2014-12-151) and the OTR Ethics Committees (41-12)); Soweto, South Africa (by the University of Witwatersrand Human Research (M130714) and the OTR Ethics Committees (1042-13 and 42-14)); and West Kiang, The Gambia (by the Gambian Government / Medical Research Council Ethics Committee (874/830)). The Jackson Heart Study (JHS) was approved by the IRB of the University of Mississippi Medical Center, Jackson State University, and Tougaloo College. All participants gave written informed consent prior to enrollment in the studies.

### Study populations

Our discovery studies included five community-based cohorts of children from sub-Saharan Africa in Kenya, Uganda, Burkina Faso, South Africa, and The Gambia. Our replication population included African American adults from the Jackson Heart Study (JHS) in Mississippi, USA. Below is a description of each of the study sites.

#### Discovery cohorts

##### Kilifi, Kenya

This included an ongoing rolling cohort evaluating malaria immunity in children in Kilifi, along the coast of Kenya.^52^ Within this cohort, children were followed-up for malaria episodes up to 8 years of age with weekly follow-ups and annual cross-sectional surveys during which anthropometry measurements, auxiliary temperature, and blood samples were taken. Iron and inflammatory markers were measured from blood samples collected at a single cross-sectional survey based on the availability of plasma samples archived at −80^0^C.

##### Entebbe, Uganda

The *Entebbe Mother and Baby Study (EMaBS)* is a prospective birth cohort that was originally designed as a randomized controlled trial to test whether anthelminthic treatment during pregnancy and early childhood was associated with differential response to vaccination or incidence of infections including malaria (http://emabs.lshtm.ac.uk/).^53^ Pregnant women from Entebbe and in their second or third trimester of pregnancy were initially enrolled at Entebbe Hospital antenatal clinic. Livebirths from these mothers were enrolled into the EMaBS study. Blood samples were collected in vacutainer tubes containing ethylenediaminetetraacetic acid (EDTA) at birth and at subsequent birthdays up to five years of age. Iron and inflammatory biomarkers were measured from a single annual visit based on the availability of stored samples.

##### Banfora, Burkina Faso

The *VAC050 ME-TRAP Malaria Vaccine Trial* recruited infants between the ages of 6 and 18 months living in the Banfora region of Burkina Faso to participate in a Phase 1/2b clinical trial of the safety, immunogenicity and efficacy of a viral-vectored prime-boost liver-stage malaria vaccine.^54^ Samples from a total of 350 infants were then selected based on suitability of samples for DNA extraction. Plasma samples were available from the infants at multiple time-points following receipt of the experimental vaccine. Samples from individuals taken at time points as close to the 12-month age as possible were prioritized for this study.

##### South Africa

*The Soweto Vaccine Response Study* consisted of infants born in Chris Hani Baragwanath Hospital living in the Soweto region of Johannesburg, South Africa who were recruited from vaccine trials^55^ that are coordinated by the Respiratory and Meningeal Pathogens Unit (http://www.rmpru.com/). Mothers of the infants were approached if their infants had received all Expanded Programme on Immunization vaccines up to six months of age. The infants were sampled prospectively at six months of age and at 12 months after receipt of measles vaccine at nine months. Single whole blood samples were collected in EDTA vacutainer tubes for measurement of iron and inflammatory markers and DNA extraction.

##### The Gambia

###### West Kiang study

All children aged two to six years (except those with chronic illness) were recruited from 10 rural villages in the West Kiang region of The Gambia during the start of a malaria season to initially investigate seasonal genetic effects of iron status, hemoglobin and haptoglobin concentrations between July and August 2001.^56^ All children had a clinical examination, anthropometric measurements, and a three-day course of mebendazole for potential hookworm infection. A blood sample was collected for measurement of iron and inflammatory markers and DNA extraction.

#### Replication cohort

##### Jackson Heart Study

This is a population-based longitudinal study of African American adults aged ≥21 years living in the Jackson, Mississippi metropolitan area in the USA.^57^ This study was designed to evaluate risks of cardiovascular disease as described elsewhere.^57,58^ Iron and inflammatory biomarkers were measured from blood samples collected at a single baseline clinic visit. Whole blood was used to extract DNA using Puregene reagents (Gentra System, Minneapolis, USA).

#### Infection GWAS

##### MalariaGEN consortium

This was a GWAS study of severe malaria in 17,000 severe malaria cases and population controls from 11 countries.^29^ For the East-Africa specific *GTF3C5* lead SNP, rs2905094, we retrieved population-specific results from all the nine African countries in the MalariaGEN database and applied fixed-effects meta-analyses by East (Kenya, Malawi, and Tanzania) and West (The Gambia, Mali, Burkina Faso, Ghana, Nigeria, and Cameroon) African sites. For the association between the hepcidin lead SNP, rs73596248 and severe malaria, we meta-analyzed all the nine African countries in the MalariaGEN database. We used the MalariaGen Kenyan GWAS to determine the association between the Kenyan specific sTfR GWAS top SNPs and severe malaria.

##### The Kenyan Bacteremia GWAS

This study comprised 1536 hospitalized Kenyan children with positive blood cultures and 2677 matched community controls as previously described.^30,31^ To determine associations between our iron-related conditionally independent SNPs and bacteremia, we identified the SNPs in the bacteremia GWAS and extracted estimates, standard errors, and p-values.

### Assays for iron and inflammatory markers

The assayed biomarkers of iron (ferritin, soluble transferrin receptors (sTfR), hepcidin, serum iron, transferrin, and unsaturated iron binding capacity (UIBC) and inflammation (C-reactive protein (CRP) and α_1_-antichymotrypsin (ACT) are shown in Supplementary Table 10 The Gambian hepcidin values were harmonized with the rest of the cohorts’ hepcidin values (measured using high sensitive DRG kit) by converting to the old DRG hepcidin assay values and then to the new high sensitive DRG hepcidin assay values.^59^ Transferrin saturation (TSAT) was calculated as (serum iron (µmol/L)/transferrin (g/L) x 25.1) x 100 or as (serum iron/ UIBC + serum iron) x 100.^60^ Since EDTA chelates iron, serum iron measurements (and TSAT) were missing in Uganda and South Africa where blood samples were collected in EDTA vacutainer tubes. Transferrin (g/L) in JHS was calculated as 0.7 x total iron binding capacity (TIBC) [µg/dL] x 0.01. Transferrin in The Gambia was similarly calculated but after calculating TIBC as UIBC + serum iron.

### Genotyping and imputation

Genotyping for DNA extracted from Uganda, Burkina Faso and South Africa was performed using the Illumina HumanOmni 2.5M-8 (‘octo’) BeadChip array version 1.1 (Illumina Inc., San Diego, USA), at the Genotyping Core facilities at the Wellcome Trust Sanger Institute. Genotyping for Kenyan and Gambian samples was performed using H3Africa Custom Genotyping Array v1.0 at the Wellcome Centre for Human Genetics, University of Oxford. JHS samples were genotyped using the Affymetrix Genome-Wide Human SNP Array 6.0. Genotypes were called from intensities using Illuminus and GenCall clustering algorithms in GenomeStudio (Illumina Inc., San Diego, USA) incorporating data from pre-determined genotypes.

We performed quality control (QC) for samples and single nucleotide polymorphisms (SNPs) using SNPs mapped to Human Genome Build 37. Using the appropriate strand files (http://www.well.ox.ac.uk/~wrayner/strand/), we removed low quality variants (those that mapped to multiple regions within the human genome or did not map to any region) and duplicate SNPs. Only autosomal SNPs were included in downstream QC analyses. We used a standardized H3A GWAS pipeline (https://github.com/h3abionet/h3agwas) to perform sample and SNP QC separately for each cohort (using identical steps).^61,62^ Samples having discordant sex information, call rates <98%, relatedness cutoffs of between 11 and 70% (accounts for cryptic relatedness), and heterozygosity <0.15 or >0.343 were removed. SNPs were retained if they passed thresholds of >99% call rate, minor allele frequency (MAF) >0.01, and Hardy–Weinberg equilibrium (HWE) adjusted p-value >0.008.

We imputed our quality-controlled genotypes using the African Genome Resources haplotype reference panel that contains 4,956 samples from 1000 Genomes Phase 3 and ∼2000 Ugandan samples and ∼100 Ethiopians/Egyptians/Namibians/South Africans (n= 93,421,145 biallelic SNPs). This reference panel is hosted by the Wellcome Sanger Institute (https://imputation.sanger.ac.uk/).^63^ We filtered out the resulting imputation dataset for SNPs with information score ≥ 0.3 and MAF ≥ 0.01 for association analyses.

### Statistical analyses

We used inverse-normal transformation to normalize the distribution of the iron biomarkers in R version 3.4.3. We performed association testing for each study site using genome-wide complex trait analysis (GCTA) software version 1.24.4^64^ fitting a linear mixed model including a genetic relatedness matrix (GRM) as a random effect. We included age and sex as covariates. Inclusion of CRP as a covariate did not change the results. Inverse normal transformation measures were used throughout our analyses and all results are reported as such. We meta-analyzed our discovery study-specific summary statistics using METASOFT version 2.0.1.^65^ Results were visualized using Manhattan and QQ plots generated using R packages *GenABEL* and *qqman*. Inflation factors (*lambda*) were calculated to check for population stratification. We used LocusZoom.js v0.12.0 (https://my.locuszoom.org/) to create GWAS regions and added LDs calculated using Plink version 1.9 from our discovery cohorts data.

Genome-wide significant SNPs (P<5×10^-8^) from discovery GWAS meta-analyses were selected for testing in the replication study of JHS. We considered a GWAS result to replicate if the effect in the replication was in the same direction as in the discovery sample, and if the association in the replication sample was statistically significant (P< 0.05).

To evaluate differences in the genetics of iron between European and African ancestry populations, we analyzed transferability of iron GWAS findings between continental African and European populations. We identified index SNPs from the largest genome-wide meta-analysis of European ancestry iron biomarkers GWAS^14,15^ for replication in our African iron GWAS. All lead/independent SNPs reported in the European ancestry iron GWAS were identified in African children with available iron GWAS data. We then compared effect sizes and P values between European and African populations. A SNP was considered to replicate if the effect in Africans was in the same direction as in the European iron GWAS, and if the association in Africans was statistically significant (P< 0.05). For each iron biomarker, we further calculated a PRS^66^ weighted using effect sizes reported in European iron biomarkers GWAS and reflecting an increase in levels of the biomarker. A linear regression model adjusted for age, sex and study site was used to determine any association between the PRS and iron biomarkers measured in African children. In Africans, we used transferrin as a proxy measure for total iron binding capacity (TIBC) that was reported in the European iron GWAS. TIBC indicates the amount of iron that can be bound to transferrin.

Since iron status might influence risk of malaria and bacterial infections,^27,28^ we investigated the association between our identified novel African-specific SNPs and these infections using the largest case-control GWAS of severe malaria^29^ and bacteremia^30,31^ as described above. Since the *SDR16C5* lead SNP appeared across all five iron GWAS African sites, we meta-analyzed severe malaria results from all nine MalariaGen African sites (The Gambia, Mali, Burkina Faso, Ghana, Nigeria, Cameroon, Malawi, Tanzania, and Kenya) using the ‘*meta’* package in R version 4.2.3 assuming fixed effects. Since the *GTF3C5* lead SNP, rs2905094, showed an East-African specific effect on transferrin levels, and was not significant in other African populations despite having a similar allele frequency, we meta-analyzed severe malaria GWAS results from East African countries only (Kenya, Malawi and Tanzania). For the *FREM3* locus lead SNP, which is Kenyan specific and very rare in other tested populations from our cohorts or in reference panels such as 1000G, we extracted its effect size in the Kenya severe malaria GWAS only.

### Functional annotation

To identify independent loci, we performed statistical fine-mapping by conducting conditional forward stepwise regression. This entailed conditioning on the lead SNP from our GWAS by treating it as an adjusting covariate and testing the remaining SNPs. Once a new independent secondary signal was identified, we performed sequential testing by adjusting for the lead SNP until no conditional tests were statistically significant.

We performed comprehensive functional annotation using FUMA (https://fuma.ctglab.nl/) to annotate our previously unreported SNPs and determine genomic risk loci. We applied an R-squared of 0.2 and used Africans (“AFR”) in the 1000G phase 3 as a reference panel. We further used VEP (https://www.ensembl.org/Tools/VEP) to annotate and prioritize variants.

To identify expression quantitative trait locus (eQTL) variants, we used GTex (https://gtexportal.org/home/) and the African Functional Genomics Resources ( https://github.com/smontgomlab/AFGR) which is the largest African eQTL database.

### Haplotype analyses

Haplotype patterns were derived from the imputed and phased genotypes using a custom R script that computed Manhattan distances followed by hierarchical clustering using the *hclust* function in R software (version 4.0.2). The resulting clusters were visualised with heat maps, and haplotype patterns were identified by visually assessing regions of homogeneity. We focused on a 72 kb region defined by 210 SNPs surrounding the rs2905094 SNP. Haplotype pattern analysis was performed separately for the ancestral and derived alleles of rs2905094, both across all countries combined and for each country individually.

### GWAS sample size and power

Power for the GWAS studies was calculated using the formulae given in Visscher et al 2017,^67^ implemented in: https://github.com/kaustubhad/gwas-power. Given an allele frequency ≥ 5% and effect size ≥ 0.33 standard deviation (SD), the overall discovery sample size of 3928 provided more than 80% power to observe true genotypic and phenotypic associations at a genome-wide significance level (type I error=5×10^-8^). For allele frequency ≥10%, we had over 80% power to see effect sizes ≥ 0.24 SD.

## Acknowledgments

We thank all study participants who contributed to this study and staff involved with consent, sample and data collection and preparation. This work was funded by Wellcome (Grant numbers [224317 to JMM], [226014 to SHA], [202800 to TNW], [103951 to AOE], and [106289 to AJM] and Grant numbers [064693, 079110, 095778 to AME]) and with core awards to the KEMRI-Wellcome Trust Research Programme (203077), The Wellcome Centre for Human Genetics (090532, 203141) and the Wellcome Sanger Institute (098051, 206194). The Jackson Heart Study (JHS) is supported and conducted in collaboration with Jackson State University (HHSN268201800013I), Tougaloo College (HHSN268201800014I), the Mississippi State Department of Health (HHSN268201800015I) and the University of Mississippi Medical Center (HHSN268201800010I, HHSN268201800011I and HHSN268201800012I) contracts from the National Heart, Lung, and Blood Institute (NHLBI) and the National Institute on Minority Health and Health Disparities (NIMHD). The authors also wish to thank the staffs and participants of the JHS. The views expressed in this manuscript are those of the authors and do not necessarily represent the views of the National Heart, Lung, and Blood Institute; the National Institutes of Health; or the U.S. Department of Health and Human Services. JJG was funded by a National Institute for Health and Care Research Clinical Lectureship. The views expressed are those of the author(s) and not necessarily those of the NIHR or the Department of Health and Social Care. This study was published with the permission of the Director of KEMRI.

## Author contributions

JMM, AJM, AME, TNW, AA, and SHA conceptualized and designed the methods for the research project; JMM performed the analyses; JMM, AJM, GB, AYC, AWM, RMM, KMA, RM, JJG, ELW, FMN, LMR, LE, ARB, SBS, SAM, AVSH, AMP, PB, GH, GDS, MSS, AME, TNW, AA, and SHA were involved in resources generation and curation of data. JMM, AJM, AME, TNW, AA, and SHA were responsible for funding acquisition. JMM and SHA wrote the manuscript and all co-authors reviewed the manuscript.

## Code availability

All analyses used preassigned code defined within software packages that are publicly available as described in Methods. Any other requests for clarifications may be sought from the corresponding authors.

## Supplementary Figures

**Supplementary Figure 1:**
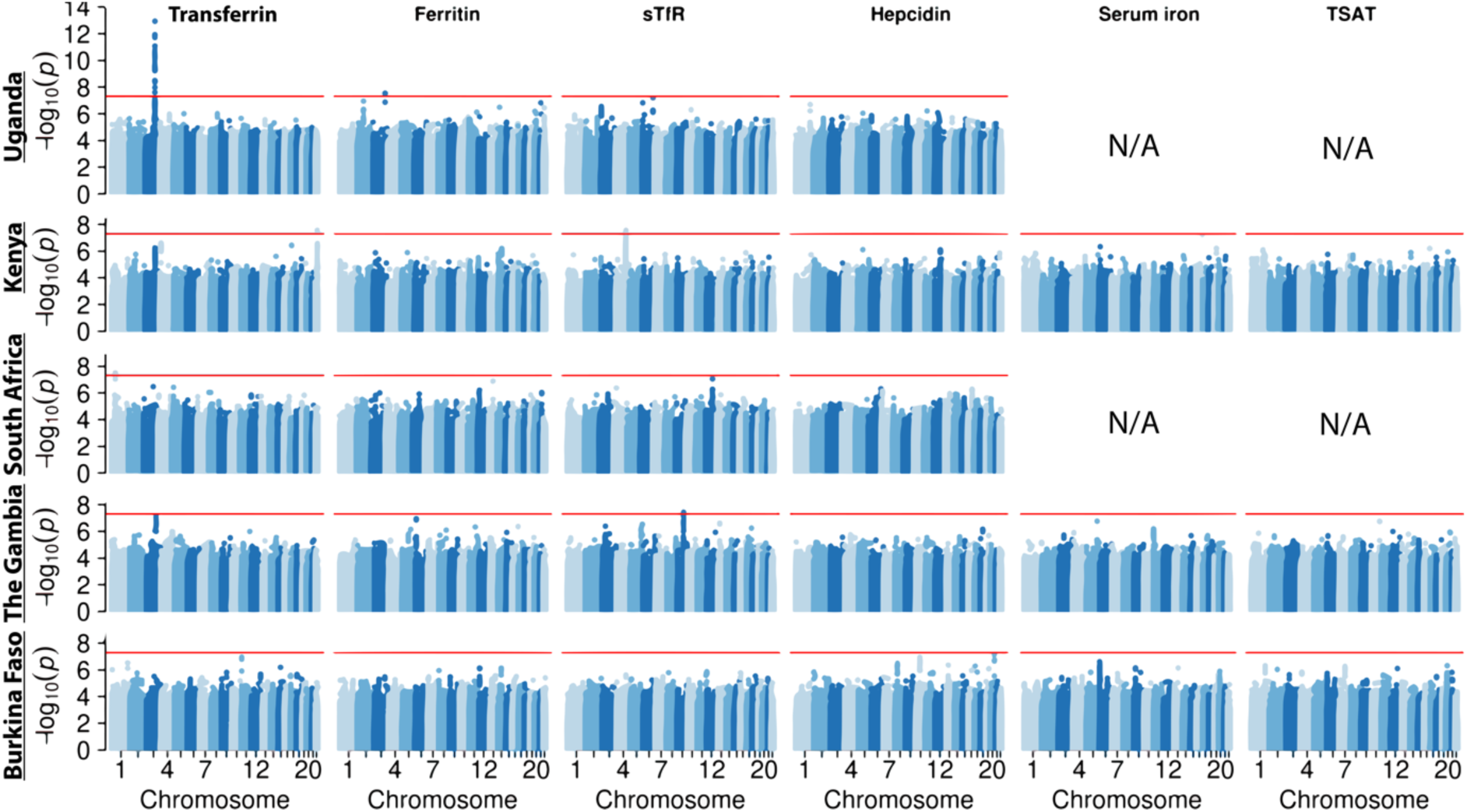
Manhattan plots for each African study site. N/A, not available.

**Supplementary Figure 2:**
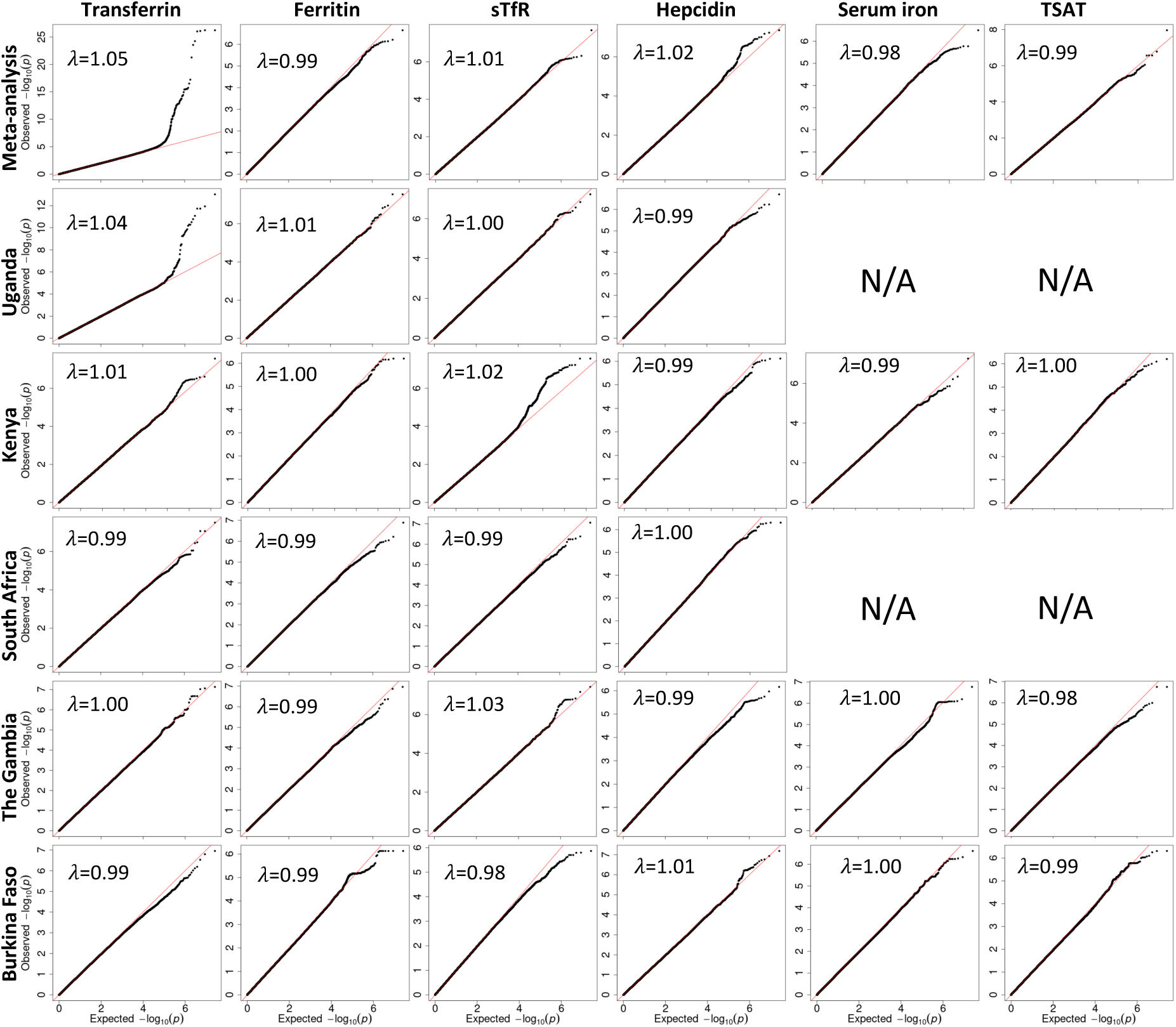
QQ plot for the discovery GWAS of iron markers.

**Supplementary Figure 3.**
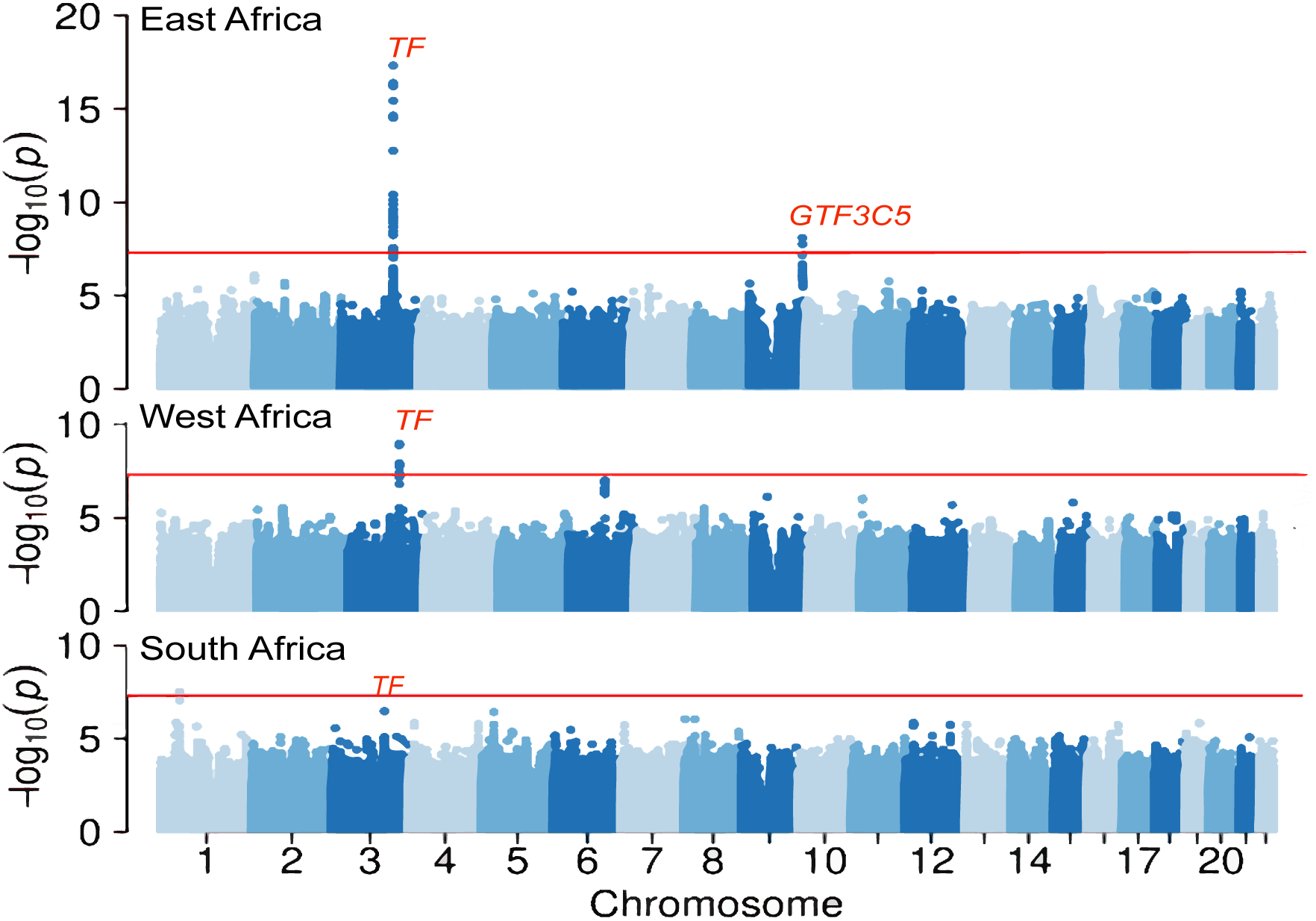
Meta-analysis of transferrin GWAS results by African region.

**Supplementary Figure 4:**
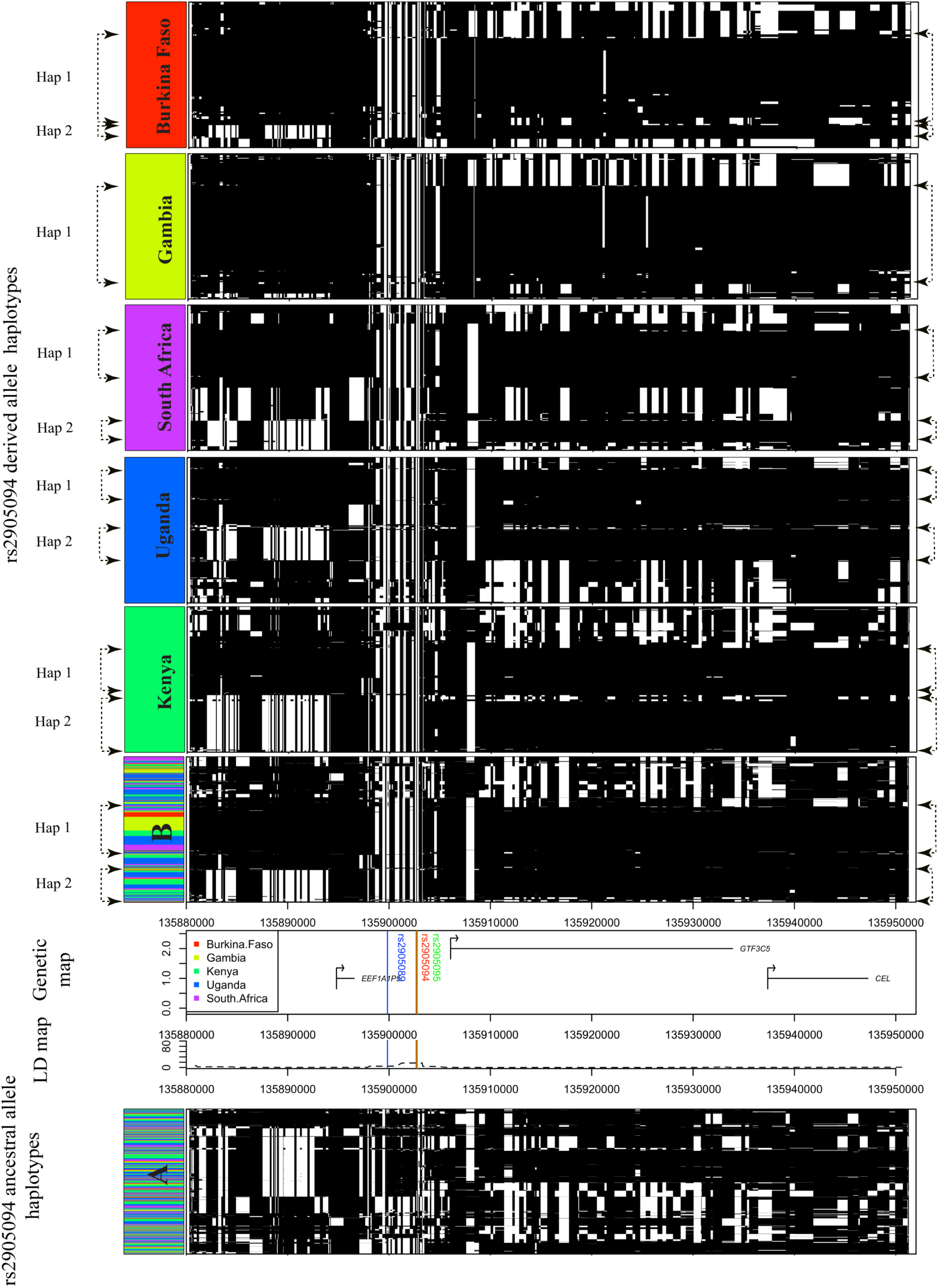
Haplotype structures, linkage disequilibrium, and genetic map of the *GTF3C5* gene region. This figure presents the haplotype structures within a 72 KB region surrounding the rs2905094 (9:135902689 (GRCh37)) allele. **Panel A** displays the structure for the ancestral allele, while **Panel B** illustrates the structure for the derived alleles across all countries combined. Additionally, haplotypes for the derived allele are shown separately for each country, as labelled in the panels. In each panel, the rows represent individual phased haplotypes, and the columns denote SNPs, sorted and clustered based on Manhattan distance. The ancestral allele is depicted by black columns, and the derived allele is represented by white columns. The dominant haplotypes identified in the mutant chromosomes are labelled as "Hap 1" and "Hap 2." The LD map is included with the Y-axis indicating the recombination rate in cM/Mb, and the X-axis displaying the chromosomal position referenced to GRCh37. The genetic map highlights the positions of three genome-wide significant SNPs - rs2905089, rs2905094, and rs2905095 - using blue, red, and green lines, respectively.

**Supplementary Figure 5.**
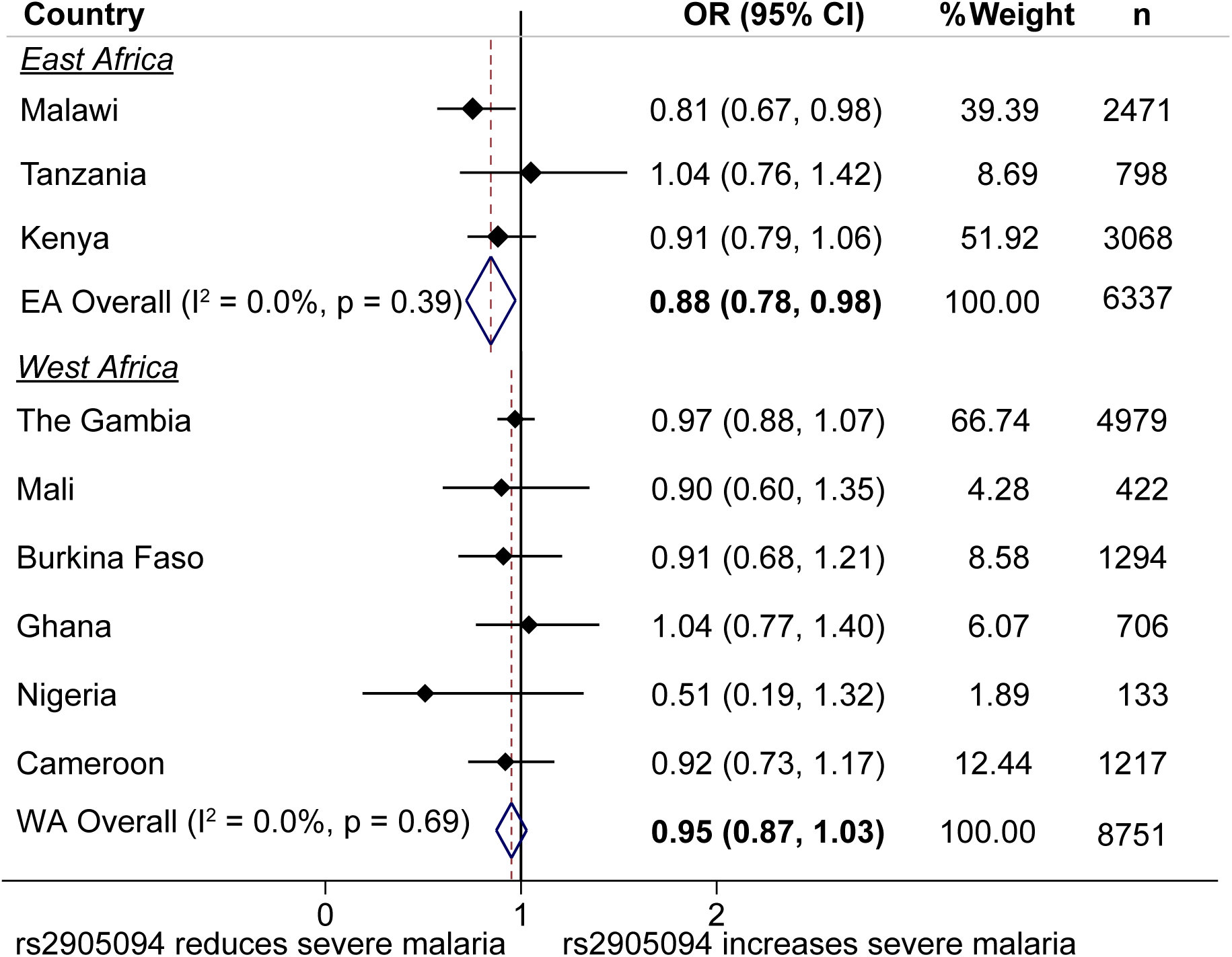
Meta-analyses of the effect sizes of the *GTF3C5* lead SNP, rs2905094 and severe malaria. We applied fixed effects meta-analysis of severe malaria GWAS results from the MalariaGen consortium by East and West African countries.

**Supplementary Figure 6.**
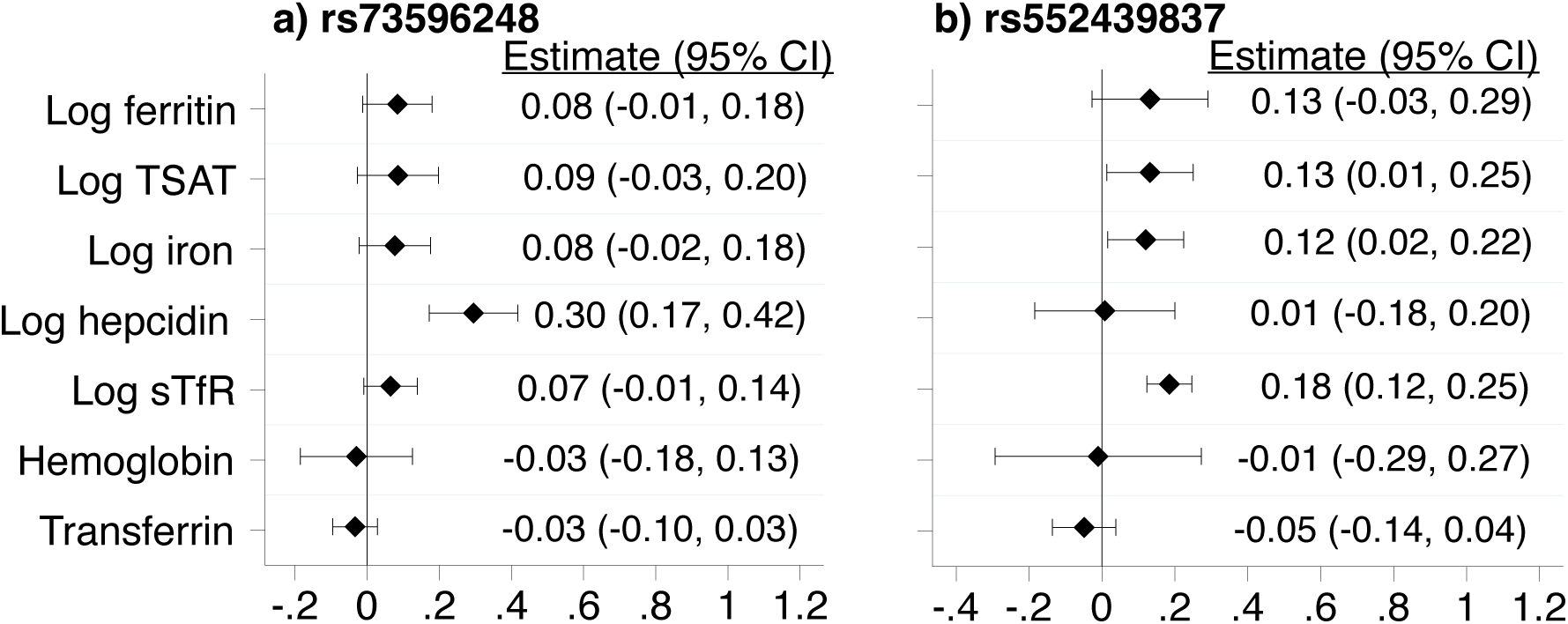
Association between **a**) hepcidin and **b**) soluble transferrin receptor GWAS lead SNPs and other iron biomarkers. Linear regression models were adjusted for age, sex, CRP, and study site. Effect sizes were derived from an additive model.

**Supplementary Figure 7.**
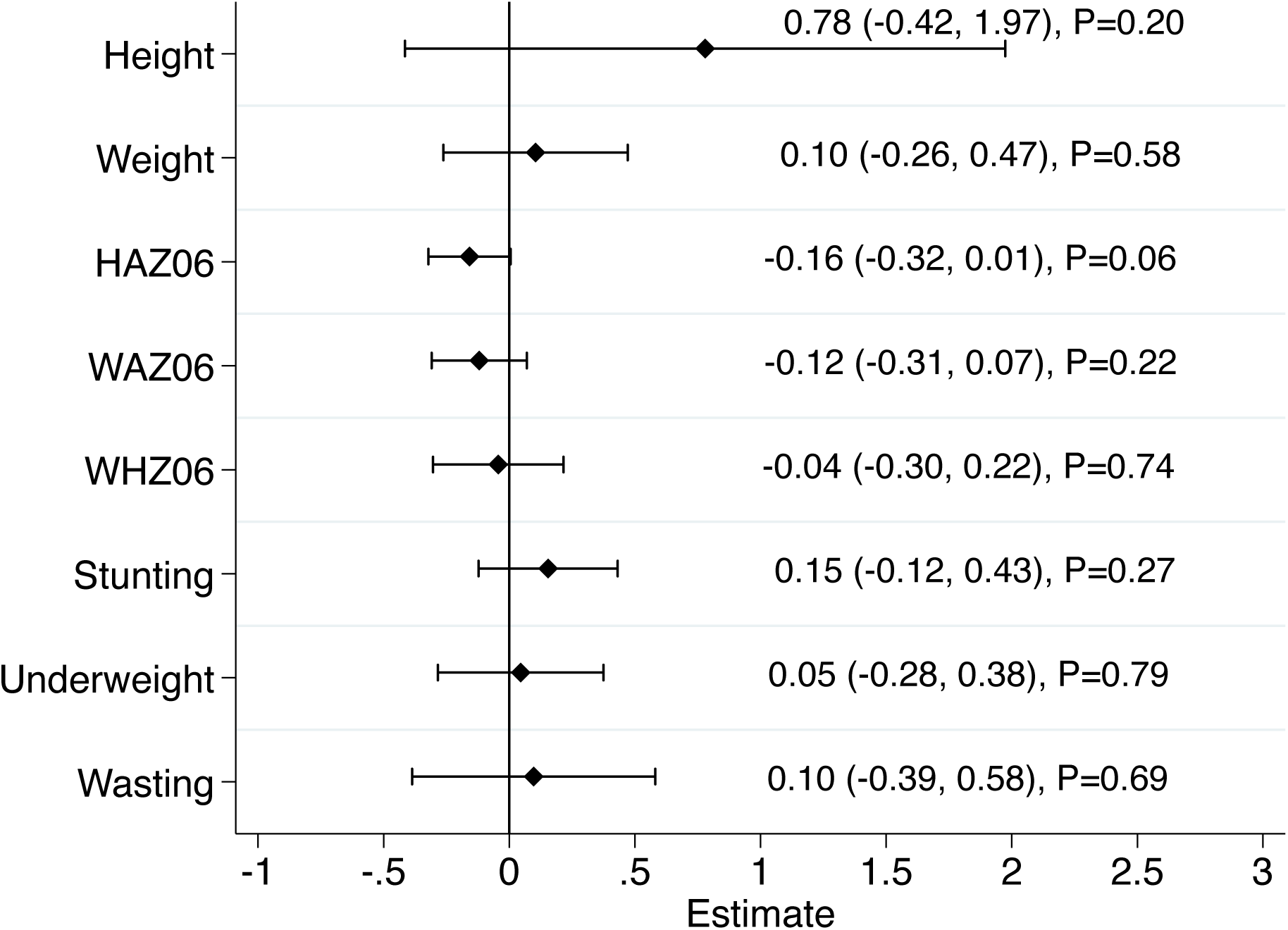
Association between hepcidin GWAS lead SNP, rs73596248 and anthropometric indices. Estimates are derived from an additive linear regression model. HAZ06, height-for-age z-score; WAZ06, weight-for-age z-score; WHZ06, weight-for-height z-score. Stunting was defined as HAZ06 < −2, underweight as WAZ06 < −2 and wasting as WHZ06 < −2 using the WHO Growth Reference Standards.

## Supplementary Tables

**Supplementary Table 1.** Characteristics of study participants.

| Characteristic | Kenya<br>n=1059 |  | Uganda<br>n=1360 |  | Burkina Faso<br>n=348 |  | South Africa<br>n=611 |  | The Gambia<br>n=550 |  | Jackson Heart Study<br>n=2868 |  |
| --- | --- | --- | --- | --- | --- | --- | --- | --- | --- | --- | --- | --- |
| Median age (IQR)<sup>a</sup> | 21.8 m (15.2-40.0) |  | 24.1 m (23.9-35.9) |  | 23.3 m (19.7-26.2) |  | 12.0 m (11.9-12.2) |  | 46.0 m (34.4-58.5) |  | 54 y (44-64) |  |
|  | n/total | % | n/total | % | n/total | % | n/total | % | n/total | % | n/total | % |
| Gender: Females | 514/1059 | 48.5 | 675/1360 | 49.6 | 173/348 | 49.7 | 294/611 | 48.1 | 250/550 | 45.5 | 1759/2868 | 61.3 |
| Inflammation<sup>b</sup> | 275/1020 | 27.0 | 314/1325 | 23.7 | 112/330 | 33.9 | 95/611 | 15.6 | 78/550 | 14.2 | 830/2864 | 29.0 |
| Malaria parasitemia<sup>c</sup> | 215/954 | 22.5 | 19/1340 | 6.8 | 66/321 | 20.6 | 0/611 | 0.0 | 59/548 | 10.8 | n/a | n/a |
| Iron deficiency<sup>d</sup> | 350/999 | 35.0 | 436/1254 | 34.8 | 115/324 | 35.5 | 251/611 | 41.1 | 118/550 | 21.5 | 298/2868 | 10.4 |
| Anemia<sup>e</sup> | 420/639 | 65.7 | 646/1299 | 49.7 | 288/331 | 87.0 | n/a | n/a | 329/545 | 60.4 | 728/2818 | 25.8 |
| Iron deficiency anemia<sup>f</sup> | 140/609 | 23.0 | 253/1197 | 21.1 | 96/309 | 31.1 | n/a | n/a | 85/545 | 15.6 | 174/2142 | 8.1 |
| Stunting<sup>g</sup> | 89/183 | 48.6 | 207/1342 | 15.4 | 109/325 | 33.5 | n/a | n/a | 167/438 | 38.1 | n/a | n/a |
| Underweight<sup>h</sup> | 95/334 | 28.4 | 112/1355 | 8.3 | 60/327 | 18.4 | n/a | n/a | 111/439 | 25.3 | n/a | n/a |
| Wasting<sup>i</sup> | 24/180 | 13.3 | 66/1340 | 4.9 | 22/325 | 6.8 | n/a | n/a | 36/438 | 8.2 | n/a | n/a |
| **Biomarker** | **n** | **GMean (SD)** | **n** | **GMean (SD)** | **n** | **GMean (SD)** | **n** | **GMean (SD)** | **n** | **GMean (SD)** | **n** | **GMean (SD)** |
| Ferritin, µg/L | 999 | 21.9 (3.1) | 1254 | 20.9 (2.9) | 324 | 22.2 (2.9) | 611 | 14.9 (2.6) | 550 | 24.4 (2.4) | 2868 | 112.2 (2.6) |
| sTfR, mg/L | 1047 | 17.8 (1.5) | 1330 | 6.7 (2.0) | 342 | 17.7 (1.7) | 610 | 10.8 (1.5) | 510 | 3.5 (1.4) | n/a | n/a |
| Hepcidin, µg/L | 976 | 5.7 (3.6) | 1321 | 6.8 (3.3) | 309 | 5.3 (4.2) | 600 | 7.8 (3.5) | 550 | 5.5 (4.7) | n/a | n/a |
| Serum iron, µmol/L | 1009 | 6.5 (2.0) | n/a | n/a | 337 | 6.0 (1.8) | n/a | n/a | 550 | 8.6 (1.6) | 2868 | 74.1 (1.5) |
| Transferrin, g/L | 998 | 2.7 (1.3) | 1320 | 2.7 (1.3) | 327 | 2.7 (1.3) | 611 | 2.6 (1.3) | 550 | 2.6 (1.2) | 2868 | 2.0 (1.2) |
| TSAT, %<sup>j</sup> | 977 | 9.4 (2.2) | n/a | n/a | 325 | 8.9 (2.0) | n/a | n/a | 550 | 12.8 (1.7) | 2868 | 25.7 (1.5) |
| Hemoglobin, g/dL | 639 | 10.2 (1.2) | 1299 | 10.8 (1.1) | 331 | 9.5 (1.1) | n/a | n/a | 545 | 10.6 (1.1) | 2818 | 12.9 (1.1) |
| Body mass index | 243 | 15.1 (1.2) | 1342 | 15.7 (1.1) | 324 | 15.4 (1.2) | n/a | n/a | 549 | 14.6 (1.1) | 2864 | 31.2 (1.2) |
| Inflammatory marker<sup>k</sup> | 1020 | 1.8 (5.2) | 1325 | 1.4 (5.1) | 330 | 2.8 (6.1) | 611 | 0.9 (5.0) | 550 | 0.4 (1.3) | 2864 | 2.5 (3.5) |

IQR, interquartile range; n/a, not available; sTfR, soluble transferrin receptors; TSAT, transferrin saturation; CRP, C-reactive protein; GMean, geometric means.
<sup>a</sup>Age in months for children and age in years for African-American (Jackson Heart Study) adults.
<sup>b</sup>Inflammation was defined as C-reactive protein > 5mg/L or α1-antichymotrypsin > 0.6g/dL (in The Gambia). Inflammatory marker values indicate CRP (mg/L) for Kenya, Uganda, Burkina Faso and South Africa, and ACT (g/dL) for The Gambia.
<sup>c</sup>Malaria parasitemia was defined as *Plasmodium falciparum* parasitemia positive at any density. Participants in South Africa and Jackson Heart Study were not exposed to malaria.
<sup>d</sup>Iron deficiency defined using WHO definition as ferritin < 12 µg/L or <30 µg/L in presence of inflammation (C-reactive protein>5mg/L or α1-antichymotrypsin >0.6g/dL) in children < 5 years or <15µg/L in children ≥5 years. In the Jackson Heart Study iron deficiency was defined as ferritin levels <30 µg/L
<sup>e</sup>Anemia was defined as hemoglobin <11 g/dL in children aged <5 years, or hemoglobin <11.5 g/dL in children ≥5 years<sup>68</sup>. In the Jackson Heart Study and Nairobi, ID was defined as ferritin <30µg/L. In the Jackson Heart Study, anemia was defined as hemoglobin <12 g/dL in women or <13 g/dL in men<sup>68,69</sup>.
<sup>f</sup>Iron deficiency anemia was defined as iron deficiency and anemia.
<sup>g</sup>Stunting was defined as height-for-age z-score < -2
<sup>h</sup>Underweight was defined as weight-for-age z-score < -2
<sup>i</sup>Wasting was defined as weight-for-height z-score < -2
<sup>j</sup>TSAT was calculated as ((Iron (µmol/L)/Transferrin (g/L) x 25.1) x 100) except in The Gambia where it was calculated using iron and unsaturated iron binding capacity. Iron and TSAT missing in Uganda and South Africa and transferrin missing in The Gambia.
<sup>k</sup>Inflammatory markers included CRP (mg/L) measured in Kenya, Uganda, Burkina Faso, South Africa, and Jackson Heart Study and ACT (g/dL) measured in The Gambia.

**Supplementary Table 2.** Summary of quality control (QC) steps for each population. The number of individuals and variants removed within each population are presented.

|  | Kenya | Uganda | Burkina Faso | South Africa | The Gambia | The Gambia | JHS |
| --- | --- | --- | --- | --- | --- | --- | --- |
| *Pre-QC* |  |  |  |  |  |  |  |
| Total number sent for genotyping | 1,312 | 1,429 | 354 | 777 | 574 | 46 | 3,021 |
| Number of variants typed and mapping to Build 37 | 2,114,814 | 2,337,579 | 2,337,579 | 2,337,579 | 2,297,939 | 2,114,814 | 874,712 |
| *Individuals QC* |  |  |  |  |  |  |  |
| Individuals with missingness >0.03 | 3 | 14 | - | 2 | 46 | - | 135 |
| Individuals with extreme heterozygosity (>0.343 or <0.15) | - | - | - | - | - | - | - |
| Individuals failing sex- check | 58 | 3 | - | 2 | 13 | - | 4 |
| Individuals IBD>0.9 and not twins / triplets | 32 | 22 | - | 24 | 9 | 2 | 3 |
| *Autosomal variants QC* |  |  |  |  |  |  |  |
| Non-autosomal variants | 36,413 | 51,101 | 51,101 | 51,101 | 36,847 | 36,413 | 34,771 |
| Autosomal duplicate variants | 28 | 9,248 | 9,248 | 9,248 | 182,365 | 28 | 2,830 |
| Autosomal variants with missingness>0.01 | 93,959 | 262,676 | 263,935 | 226,503 | 113,215 | 89,922 | 122,182 |
| Autosomal variants with MAF<0.01 | 211,678 | 392,392 | 399,486 | 416,608 | 222,631 | 231,339 | 342 |
| Autosomal variants in Hardy-Weinberg disequilibrium<br>(P<0.008) | 29,735 | 29,266 | 20,089 | 25,174 | 21,989 | 6,544 | 17,258 |
| *Post-QC* |  |  |  |  |  |  |  |
| Total individuals remaining | 1,219 | 1,390 | 354 | 749 | 506 | 44 | 2,879 |
| Total autosomal variants remaining | 1,743,001 | 1,592,896 | 1,593,720 | 1,608,945 | 1,720,892 | 1,750,568 | 697,329 |

Genotyping for the 574 and 46 Gambian samples was done separately and then merged after quality control giving total individual remaining=550 and total autosomal variants remaining 1,776,801. JHS, Jackson Heart Study.

**Supplementary Table 3.** Number of variants imputed into individual datasets.

| **Cohort** | **Total imputed** | **Total high quality** | **Total tested for association** |
| --- | --- | --- | --- |
| Kenya | 89,838,087 | 13,870,649 | 7,756,141 |
| Uganda | 89,838,087 | 13,982,934 | 13,765,898 |
| Burkina Faso | 89,838,087 | 13,128,631 | 13,042,768 |
| South Africa | 89,838,087 | 15,010,497 | 14,457,588 |
| The Gambia | 89,838,087 | 12,770,741 | 10,295,094 |
| Jackson Heart Study | 89,838,087 | 13,455,566 | 13,349,077 |

High quality SNPs are those only with an information score greater than 0.3 and those tested for association were only those of high quality with a minor allele frequency (MAF) greater than 0.01.

**Supplementary Table 4.**
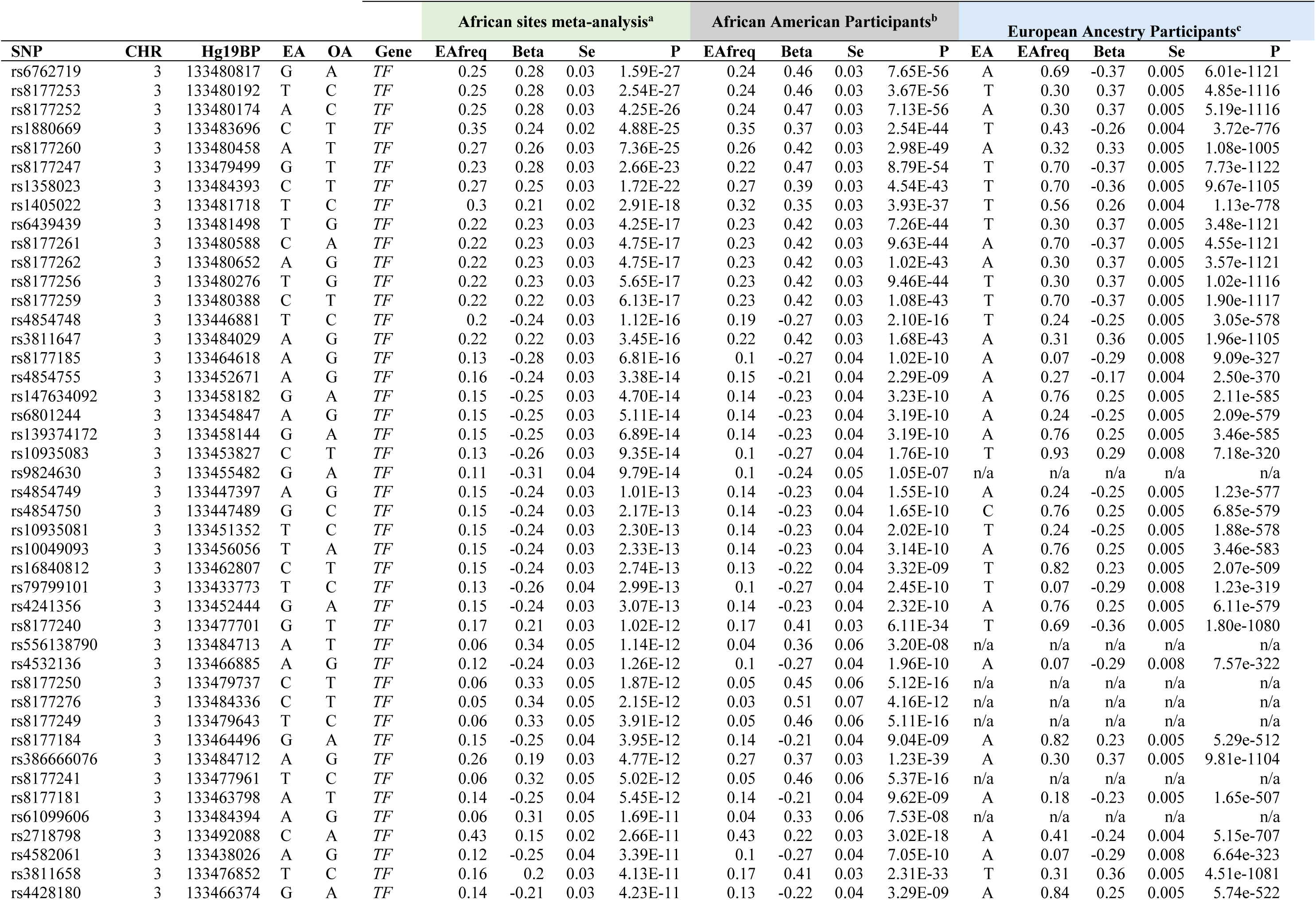
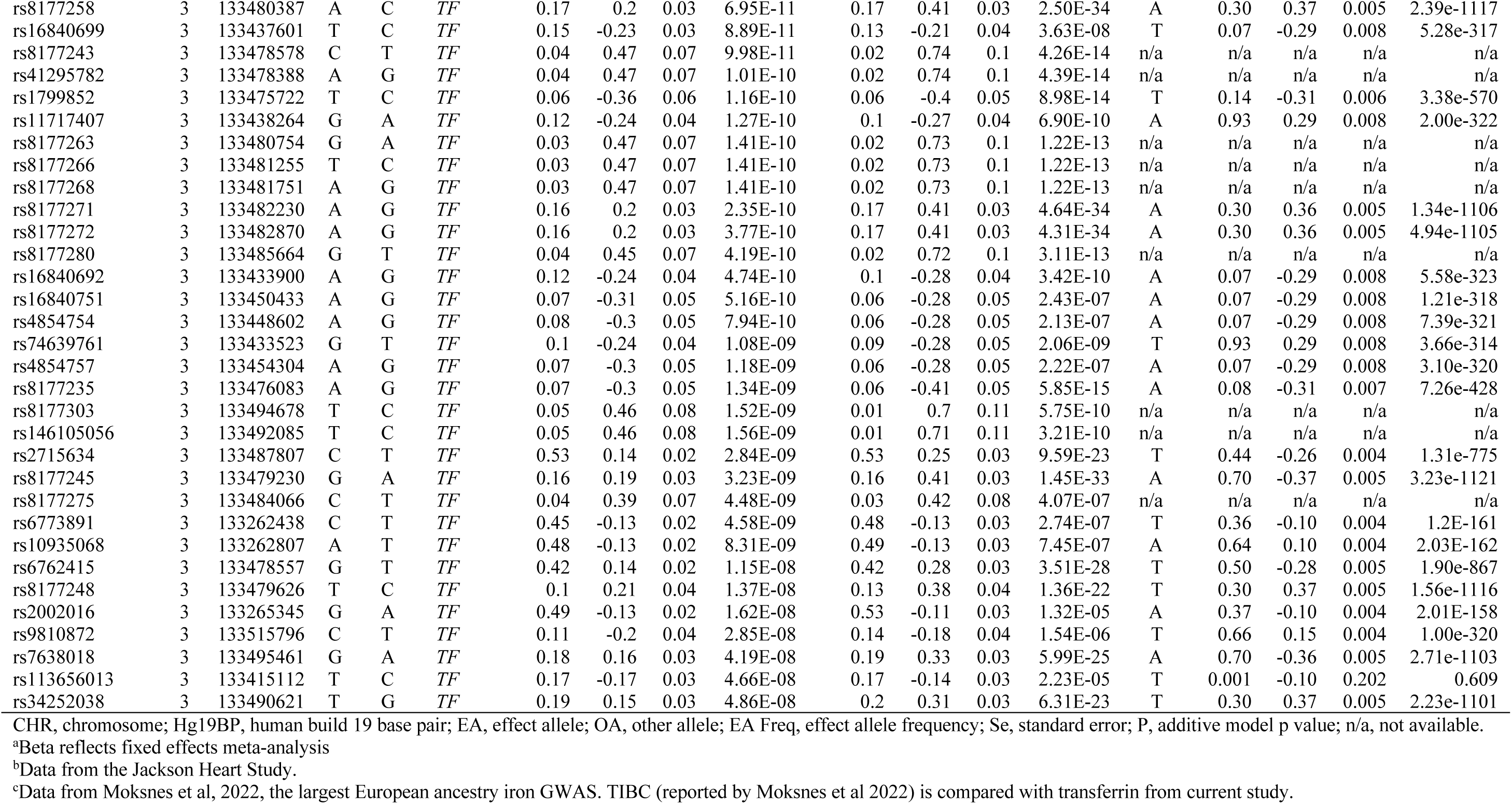
Genome-wide significant variants associated with transferrin levels at the *TF* locus in a meta-analysis of African sites, in African American and European populations.

| SNP | CHR | Hg19BP | EA | OA | Gene | African sites meta-analysis<sup>a</sup> |  |  |  | African American Participants<sup>b</sup> |  |  |  | European Ancestry Participants<sup>c</sup> |  |  |  |  |
| --- | --- | --- | --- | --- | --- | --- | --- | --- | --- | --- | --- | --- | --- | --- | --- | --- | --- | --- |
|  |  |  |  |  |  | Eafreq | Beta | Se | P | Eafreq | Beta | Se | P | EA | Eafreq | Beta | Se | P |
| rs6762719 | 3 | 133480817 | G | A | *TF* | 0.25 | 0.28 | 0.03 | 1.59E-27 | 0.24 | 0.46 | 0.03 | 7.65E-56 | A | 0.69 | -0.37 | 0.005 | 6.01e-1121 |
| rs8177253 | 3 | 133480192 | T | C | *TF* | 0.25 | 0.28 | 0.03 | 2.54E-27 | 0.24 | 0.46 | 0.03 | 3.67E-56 | T | 0.30 | 0.37 | 0.005 | 4.85e-1116 |
| rs8177252 | 3 | 133480174 | A | C | *TF* | 0.25 | 0.28 | 0.03 | 4.25E-26 | 0.24 | 0.47 | 0.03 | 7.13E-56 | A | 0.30 | 0.37 | 0.005 | 5.19e-1116 |
| rs1880669 | 3 | 133483696 | C | T | *TF* | 0.35 | 0.24 | 0.02 | 4.88E-25 | 0.35 | 0.37 | 0.03 | 2.54E-44 | T | 0.43 | -0.26 | 0.004 | 3.72e-776 |
| rs8177260 | 3 | 133480458 | A | T | *TF* | 0.27 | 0.26 | 0.03 | 7.36E-25 | 0.26 | 0.42 | 0.03 | 2.98E-49 | A | 0.32 | 0.33 | 0.005 | 1.08e-1005 |
| rs8177247 | 3 | 133479499 | G | T | *TF* | 0.23 | 0.28 | 0.03 | 2.66E-23 | 0.22 | 0.47 | 0.03 | 8.79E-54 | T | 0.70 | -0.37 | 0.005 | 7.73e-1122 |
| rs1358023 | 3 | 133484393 | C | T | *TF* | 0.27 | 0.25 | 0.03 | 1.72E-22 | 0.27 | 0.39 | 0.03 | 4.54E-43 | T | 0.70 | -0.36 | 0.005 | 9.67e-1105 |
| rs1405022 | 3 | 133481718 | T | C | *TF* | 0.3 | 0.21 | 0.02 | 2.91E-18 | 0.32 | 0.35 | 0.03 | 3.93E-37 | T | 0.56 | 0.26 | 0.004 | 1.13e-778 |
| rs6439439 | 3 | 133481498 | T | G | *TF* | 0.22 | 0.23 | 0.03 | 4.25E-17 | 0.23 | 0.42 | 0.03 | 7.26E-44 | T | 0.30 | 0.37 | 0.005 | 3.48e-1121 |
| rs8177261 | 3 | 133480588 | C | A | *TF* | 0.22 | 0.23 | 0.03 | 4.75E-17 | 0.23 | 0.42 | 0.03 | 9.63E-44 | A | 0.70 | -0.37 | 0.005 | 4.55e-1121 |
| rs8177262 | 3 | 133480652 | A | G | *TF* | 0.22 | 0.23 | 0.03 | 4.75E-17 | 0.23 | 0.42 | 0.03 | 1.02E-43 | A | 0.30 | 0.37 | 0.005 | 3.57e-1121 |
| rs8177256 | 3 | 133480276 | T | G | *TF* | 0.22 | 0.23 | 0.03 | 5.65E-17 | 0.23 | 0.42 | 0.03 | 9.46E-44 | T | 0.30 | 0.37 | 0.005 | 1.02e-1116 |
| rs8177259 | 3 | 133480388 | C | T | *TF* | 0.22 | 0.22 | 0.03 | 6.13E-17 | 0.23 | 0.42 | 0.03 | 1.08E-43 | T | 0.70 | -0.37 | 0.005 | 1.90e-1117 |
| rs4854748 | 3 | 133446881 | T | C | *TF* | 0.2 | -0.24 | 0.03 | 1.12E-16 | 0.19 | -0.27 | 0.03 | 2.10E-16 | T | 0.24 | -0.25 | 0.005 | 3.05e-578 |
| rs3811647 | 3 | 133484029 | A | G | *TF* | 0.22 | 0.22 | 0.03 | 3.45E-16 | 0.22 | 0.42 | 0.03 | 1.68E-43 | A | 0.31 | 0.36 | 0.005 | 1.96e-1105 |
| rs8177185 | 3 | 133464618 | A | G | *TF* | 0.13 | -0.28 | 0.03 | 6.81E-16 | 0.1 | -0.27 | 0.04 | 1.02E-10 | A | 0.07 | -0.29 | 0.008 | 9.09e-327 |
| rs4854755 | 3 | 133452671 | A | G | *TF* | 0.16 | -0.24 | 0.03 | 3.38E-14 | 0.15 | -0.21 | 0.04 | 2.29E-09 | A | 0.27 | -0.17 | 0.004 | 2.50e-370 |
| rs147634092 | 3 | 133458182 | G | A | *TF* | 0.15 | -0.25 | 0.03 | 4.70E-14 | 0.14 | -0.23 | 0.04 | 3.23E-10 | A | 0.76 | 0.25 | 0.005 | 2.11e-585 |
| rs6801244 | 3 | 133454847 | A | G | *TF* | 0.15 | -0.25 | 0.03 | 5.11E-14 | 0.14 | -0.23 | 0.04 | 3.19E-10 | A | 0.24 | -0.25 | 0.005 | 2.09e-579 |
| rs139374172 | 3 | 133458144 | G | A | *TF* | 0.15 | -0.25 | 0.03 | 6.89E-14 | 0.14 | -0.23 | 0.04 | 3.19E-10 | A | 0.76 | 0.25 | 0.005 | 3.46e-585 |
| rs10935083 | 3 | 133453827 | C | T | *TF* | 0.13 | -0.26 | 0.03 | 9.35E-14 | 0.1 | -0.27 | 0.04 | 1.76E-10 | T | 0.93 | 0.29 | 0.008 | 7.18e-320 |
| rs9824630 | 3 | 133455482 | G | A | *TF* | 0.11 | -0.31 | 0.04 | 9.79E-14 | 0.1 | -0.24 | 0.05 | 1.05E-07 | n/a | n/a | n/a | n/a | n/a |
| rs4854749 | 3 | 133447397 | A | G | *TF* | 0.15 | -0.24 | 0.03 | 1.01E-13 | 0.14 | -0.23 | 0.04 | 1.55E-10 | A | 0.24 | -0.25 | 0.005 | 1.23e-577 |
| rs4854750 | 3 | 133447489 | G | C | *TF* | 0.15 | -0.24 | 0.03 | 2.17E-13 | 0.14 | -0.23 | 0.04 | 1.65E-10 | C | 0.76 | 0.25 | 0.005 | 6.85e-579 |
| rs10935081 | 3 | 133451352 | T | C | *TF* | 0.15 | -0.24 | 0.03 | 2.30E-13 | 0.14 | -0.23 | 0.04 | 2.02E-10 | T | 0.24 | -0.25 | 0.005 | 1.88e-578 |
| rs10049093 | 3 | 133456056 | T | A | *TF* | 0.15 | -0.24 | 0.03 | 2.33E-13 | 0.14 | -0.23 | 0.04 | 3.14E-10 | A | 0.76 | 0.25 | 0.005 | 3.46e-583 |
| rs16840812 | 3 | 133462807 | C | T | *TF* | 0.15 | -0.24 | 0.03 | 2.74E-13 | 0.13 | -0.22 | 0.04 | 3.32E-09 | T | 0.82 | 0.23 | 0.005 | 2.07e-509 |
| rs79799101 | 3 | 133433773 | T | C | *TF* | 0.13 | -0.26 | 0.04 | 2.99E-13 | 0.1 | -0.27 | 0.04 | 2.45E-10 | T | 0.07 | -0.29 | 0.008 | 1.23e-319 |
| rs4241356 | 3 | 133452444 | G | A | *TF* | 0.15 | -0.24 | 0.03 | 3.07E-13 | 0.14 | -0.23 | 0.04 | 2.32E-10 | A | 0.76 | 0.25 | 0.005 | 6.11e-579 |
| rs8177240 | 3 | 133477701 | G | T | *TF* | 0.17 | 0.21 | 0.03 | 1.02E-12 | 0.17 | 0.41 | 0.03 | 6.11E-34 | T | 0.69 | -0.36 | 0.005 | 1.80e-1080 |
| rs556138790 | 3 | 133484713 | A | T | *TF* | 0.06 | 0.34 | 0.05 | 1.14E-12 | 0.04 | 0.36 | 0.06 | 3.20E-08 | n/a | n/a | n/a | n/a | n/a |
| rs4532136 | 3 | 133466885 | A | G | *TF* | 0.12 | -0.24 | 0.03 | 1.26E-12 | 0.1 | -0.27 | 0.04 | 1.96E-10 | A | 0.07 | -0.29 | 0.008 | 7.57e-322 |
| rs8177250 | 3 | 133479737 | C | T | *TF* | 0.06 | 0.33 | 0.05 | 1.87E-12 | 0.05 | 0.45 | 0.06 | 5.12E-16 | n/a | n/a | n/a | n/a | n/a |
| rs8177276 | 3 | 133484336 | C | T | *TF* | 0.05 | 0.34 | 0.05 | 2.15E-12 | 0.03 | 0.51 | 0.07 | 4.16E-12 | n/a | n/a | n/a | n/a | n/a |
| rs8177249 | 3 | 133479643 | T | C | *TF* | 0.06 | 0.33 | 0.05 | 3.91E-12 | 0.05 | 0.46 | 0.06 | 5.11E-16 | n/a | n/a | n/a | n/a | n/a |
| rs8177184 | 3 | 133464496 | G | A | *TF* | 0.15 | -0.25 | 0.04 | 3.95E-12 | 0.14 | -0.21 | 0.04 | 9.04E-09 | A | 0.82 | 0.23 | 0.005 | 5.29e-512 |
| rs386666076 | 3 | 133484712 | A | G | *TF* | 0.26 | 0.19 | 0.03 | 4.77E-12 | 0.27 | 0.37 | 0.03 | 1.23E-39 | A | 0.30 | 0.37 | 0.005 | 9.81e-1104 |
| rs8177241 | 3 | 133477961 | T | C | *TF* | 0.06 | 0.32 | 0.05 | 5.02E-12 | 0.05 | 0.46 | 0.06 | 5.37E-16 | n/a | n/a | n/a | n/a | n/a |
| rs8177181 | 3 | 133463798 | A | T | *TF* | 0.14 | -0.25 | 0.04 | 5.45E-12 | 0.14 | -0.21 | 0.04 | 9.62E-09 | A | 0.18 | -0.23 | 0.005 | 1.65e-507 |
| rs61099606 | 3 | 133484394 | A | G | *TF* | 0.06 | 0.31 | 0.05 | 1.69E-11 | 0.04 | 0.33 | 0.06 | 7.53E-08 | n/a | n/a | n/a | n/a | n/a |
| rs2718798 | 3 | 133492088 | C | A | *TF* | 0.43 | 0.15 | 0.02 | 2.66E-11 | 0.43 | 0.22 | 0.03 | 3.02E-18 | A | 0.41 | -0.24 | 0.004 | 5.15e-707 |
| rs4582061 | 3 | 133438026 | A | G | *TF* | 0.12 | -0.25 | 0.04 | 3.39E-11 | 0.1 | -0.27 | 0.04 | 7.05E-10 | A | 0.07 | -0.29 | 0.008 | 6.64e-323 |
| rs3811658 | 3 | 133476852 | T | C | *TF* | 0.16 | 0.2 | 0.03 | 4.13E-11 | 0.17 | 0.41 | 0.03 | 2.31E-33 | T | 0.31 | 0.36 | 0.005 | 4.51e-1081 |
| rs4428180 | 3 | 133466374 | G | A | *TF* | 0.14 | -0.21 | 0.03 | 4.23E-11 | 0.13 | -0.22 | 0.04 | 3.29E-09 | A | 0.84 | 0.25 | 0.005 | 5.74e-522 |
| rs8177258 | 3 | 133480387 | A | C | TF | 0.17 | 0.2 | 0.03 | 6.95E-11 | 0.17 | 0.41 | 0.03 | 2.50E-34 | A | 0.30 | 0.37 | 0.005 | 2.39e-1117 |
| rs16840699 | 3 | 133437601 | T | C | TF | 0.15 | -0.23 | 0.03 | 8.89E-11 | 0.13 | -0.21 | 0.04 | 3.63E-08 | T | 0.07 | -0.29 | 0.008 | 5.28e-317 |
| rs8177243 | 3 | 133478578 | C | T | TF | 0.04 | 0.47 | 0.07 | 9.98E-11 | 0.02 | 0.74 | 0.1 | 4.26E-14 | n/a | n/a | n/a | n/a | n/a |
| rs41295782 | 3 | 133478388 | A | G | TF | 0.04 | 0.47 | 0.07 | 1.01E-10 | 0.02 | 0.74 | 0.1 | 4.39E-14 | n/a | n/a | n/a | n/a | n/a |
| rs1799852 | 3 | 133475722 | T | C | TF | 0.06 | -0.36 | 0.06 | 1.16E-10 | 0.06 | -0.4 | 0.05 | 8.98E-14 | T | 0.14 | -0.31 | 0.006 | 3.38e-570 |
| rs11717407 | 3 | 133438264 | G | A | TF | 0.12 | -0.24 | 0.04 | 1.27E-10 | 0.1 | -0.27 | 0.04 | 6.90E-10 | A | 0.93 | 0.29 | 0.008 | 2.00e-322 |
| rs8177263 | 3 | 133480754 | G | A | TF | 0.03 | 0.47 | 0.07 | 1.41E-10 | 0.02 | 0.73 | 0.1 | 1.22E-13 | n/a | n/a | n/a | n/a | n/a |
| rs8177266 | 3 | 133481255 | T | C | TF | 0.03 | 0.47 | 0.07 | 1.41E-10 | 0.02 | 0.73 | 0.1 | 1.22E-13 | n/a | n/a | n/a | n/a | n/a |
| rs8177268 | 3 | 133481751 | A | G | TF | 0.03 | 0.47 | 0.07 | 1.41E-10 | 0.02 | 0.73 | 0.1 | 1.22E-13 | n/a | n/a | n/a | n/a | n/a |
| rs8177271 | 3 | 133482230 | A | G | TF | 0.16 | 0.2 | 0.03 | 2.35E-10 | 0.17 | 0.41 | 0.03 | 4.64E-34 | A | 0.30 | 0.36 | 0.005 | 1.34e-1106 |
| rs8177272 | 3 | 133482870 | A | G | TF | 0.16 | 0.2 | 0.03 | 3.77E-10 | 0.17 | 0.41 | 0.03 | 4.31E-34 | A | 0.30 | 0.36 | 0.005 | 4.94e-1105 |
| rs8177280 | 3 | 133485664 | G | T | TF | 0.04 | 0.45 | 0.07 | 4.19E-10 | 0.02 | 0.72 | 0.1 | 3.11E-13 | n/a | n/a | n/a | n/a | n/a |
| rs16840692 | 3 | 133433900 | A | G | TF | 0.12 | -0.24 | 0.04 | 4.74E-10 | 0.1 | -0.28 | 0.04 | 3.42E-10 | A | 0.07 | -0.29 | 0.008 | 5.58e-323 |
| rs16840751 | 3 | 133450433 | A | G | TF | 0.07 | -0.31 | 0.05 | 5.16E-10 | 0.06 | -0.28 | 0.05 | 2.43E-07 | A | 0.07 | -0.29 | 0.008 | 1.21e-318 |
| rs4854754 | 3 | 133448602 | A | G | TF | 0.08 | -0.3 | 0.05 | 7.94E-10 | 0.06 | -0.28 | 0.05 | 2.13E-07 | A | 0.07 | -0.29 | 0.008 | 7.39e-321 |
| rs74639761 | 3 | 133433523 | G | T | TF | 0.1 | -0.24 | 0.04 | 1.08E-09 | 0.09 | -0.28 | 0.05 | 2.06E-09 | T | 0.93 | 0.29 | 0.008 | 3.66e-314 |
| rs4854757 | 3 | 133454304 | A | G | TF | 0.07 | -0.3 | 0.05 | 1.18E-09 | 0.06 | -0.28 | 0.05 | 2.22E-07 | A | 0.07 | -0.29 | 0.008 | 3.10e-320 |
| rs8177235 | 3 | 133476083 | A | G | TF | 0.07 | -0.3 | 0.05 | 1.34E-09 | 0.06 | -0.41 | 0.05 | 5.85E-15 | A | 0.08 | -0.31 | 0.007 | 7.26e-428 |
| rs8177303 | 3 | 133494678 | T | C | TF | 0.05 | 0.46 | 0.08 | 1.52E-09 | 0.01 | 0.7 | 0.11 | 5.75E-10 | n/a | n/a | n/a | n/a | n/a |
| rs146105056 | 3 | 133492085 | T | C | TF | 0.05 | 0.46 | 0.08 | 1.56E-09 | 0.01 | 0.71 | 0.11 | 3.21E-10 | n/a | n/a | n/a | n/a | n/a |
| rs2715634 | 3 | 133487807 | C | T | TF | 0.53 | 0.14 | 0.02 | 2.84E-09 | 0.53 | 0.25 | 0.03 | 9.59E-23 | T | 0.44 | -0.26 | 0.004 | 1.31e-775 |
| rs8177245 | 3 | 133479230 | G | A | TF | 0.16 | 0.19 | 0.03 | 3.23E-09 | 0.16 | 0.41 | 0.03 | 1.45E-33 | A | 0.70 | -0.37 | 0.005 | 3.23e-1121 |
| rs8177275 | 3 | 133484066 | C | T | TF | 0.04 | 0.39 | 0.07 | 4.48E-09 | 0.03 | 0.42 | 0.08 | 4.07E-07 | n/a | n/a | n/a | n/a | n/a |
| rs6773891 | 3 | 133262438 | C | T | TF | 0.45 | -0.13 | 0.02 | 4.58E-09 | 0.48 | -0.13 | 0.03 | 2.74E-07 | T | 0.36 | -0.10 | 0.004 | 1.2E-161 |
| rs10935068 | 3 | 133262807 | A | T | TF | 0.48 | -0.13 | 0.02 | 8.31E-09 | 0.49 | -0.13 | 0.03 | 7.45E-07 | A | 0.64 | 0.10 | 0.004 | 2.03E-162 |
| rs6762415 | 3 | 133478557 | G | T | TF | 0.42 | 0.14 | 0.02 | 1.15E-08 | 0.42 | 0.28 | 0.03 | 3.51E-28 | T | 0.50 | -0.28 | 0.005 | 1.90e-867 |
| rs8177248 | 3 | 133479626 | T | C | TF | 0.1 | 0.21 | 0.04 | 1.37E-08 | 0.13 | 0.38 | 0.04 | 1.36E-22 | T | 0.30 | 0.37 | 0.005 | 1.56e-1116 |
| rs2002016 | 3 | 133265345 | G | A | TF | 0.49 | -0.13 | 0.02 | 1.62E-08 | 0.53 | -0.11 | 0.03 | 1.32E-05 | A | 0.37 | -0.10 | 0.004 | 2.01E-158 |
| rs9810872 | 3 | 133515796 | C | T | TF | 0.11 | -0.2 | 0.04 | 2.85E-08 | 0.14 | -0.18 | 0.04 | 1.54E-06 | T | 0.66 | 0.15 | 0.004 | 1.00e-320 |
| rs7638018 | 3 | 133495461 | G | A | TF | 0.18 | 0.16 | 0.03 | 4.19E-08 | 0.19 | 0.33 | 0.03 | 5.99E-25 | A | 0.70 | -0.36 | 0.005 | 2.71e-1103 |
| rs113656013 | 3 | 133415112 | T | C | TF | 0.17 | -0.17 | 0.03 | 4.66E-08 | 0.17 | -0.14 | 0.03 | 2.23E-05 | T | 0.001 | -0.10 | 0.202 | 0.609 |
| rs34252038 | 3 | 133490621 | T | G | TF | 0.19 | 0.15 | 0.03 | 4.86E-08 | 0.2 | 0.31 | 0.03 | 6.31E-23 | T | 0.30 | 0.37 | 0.005 | 2.23e-1101 |

CHR, chromosome; Hg19BP, human build 19 base pair; EA, effect allele; OA, other allele; EA Freq, effect allele frequency; Se, standard error; P, additive model p value; n/a, not available.
<sup>a</sup>Beta reflects fixed effects meta-analysis
<sup>b</sup>Data from the Jackson Heart Study.
<sup>c</sup>Data from Moksnes et al, 2022, the largest European ancestry iron GWAS. TIBC (reported by Moksnes et al 2022) is compared with transferrin from current study.

**Supplementary Table 5.** Genome-wide significant SNPs associated with transferrin levels at *GTF3C5* locus in meta-analysis of Kenya and Uganda and how they compare with other populations.

|  | SNP | rs2905094 | rs2905095 | rs2905089 |
| --- | --- | --- | --- | --- |
|  | CHR | 9 | 9 | 9 |
|  | Hg19BP | 135902689 | 135902793 | 135899844 |
|  | EA | T | T | C |
|  | OA | C | C | A |
| Kenya+Uganda meta-analysis | EAFreq | 0.16 | 0.18 | 0.25 |
|  | Beta | -0.23 | -0.22 | -0.18 |
|  | Se | 0.04 | 0.04 | 0.03 |
|  | P | 8.08E-09 | 1.75E-08 | 6.74E-08 |
| Uganda | EAFreq | 0.18 | 0.20 | 0.29 |
|  | Beta | -0.20 | -0.19 | -0.18 |
|  | Se | 0.05 | 0.05 | 0.04 |
|  | P | 9.82E-05 | 9.39E-05 | 3.60E-05 |
| Kenya | EAFreq | 0.14 | 0.16 | 0.21 |
|  | Beta | -0.29 | -0.26 | -0.20 |
|  | Se | 0.07 | 0.06 | 0.06 |
|  | P | 1.15E-05 | 3.17E-05 | 4.94E-04 |
| South Africa | EAFreq | 0.14 | 0.16 | 0.25 |
|  | Beta | -0.15 | -0.13 | -0.10 |
|  | Se | 0.08 | 0.08 | 0.07 |
|  | P | 7.35E-02 | 9.95E-02 | 1.59E-01 |
| The Gambia | EAFreq | 0.23 | 0.25 | 0.32 |
|  | Beta | 0.12 | 0.06 | -0.04 |
|  | Se | 0.07 | 0.07 | 0.06 |
|  | P | 1.06E-01 | 3.55E-01 | 5.81E-01 |
| Burkina Faso | EAFreq | 0.17 | 0.18 | 0.25 |
|  | Beta | -0.03 | -0.06 | 0.09 |
|  | Se | 0.10 | 0.09 | 0.09 |
|  | P | 7.86E-01 | 5.08E-01 | 3.02E-01 |
| Jackson Heart Study | EAFreq | 0.18 | 0.19 | 0.27 |
|  | Beta | -0.02 | -0.03 | -0.02 |
|  | Se | 0.04 | 0.04 | 0.03 |
|  | P | 5.78E-01 | 4.62E-01 | 5.43E-01 |
| European Ancestry Meta-analysis<sup>a</sup> | EAFreq | 0.33 | 0.30 | 0.33 |
|  | Beta | 0.004 | 0.01 | 0.001 |
|  | Se | 0.004 | 0.007 | 0.004 |
|  | P | 3.20E-01 | 1.10E-01 | 7.40E-01 |

CHR, chromosome; Hg19BP, human build 19 base pair; EA, effect allele; OA, other allele; EA Freq, effect allele frequency; Se, standard error; P, additive model p value
<sup>a</sup>Effect of SNPs on transferrin iron binding capacity reported by Moksnes et al 2022 in European ancestry participants.

**Supplementary Table 6.**
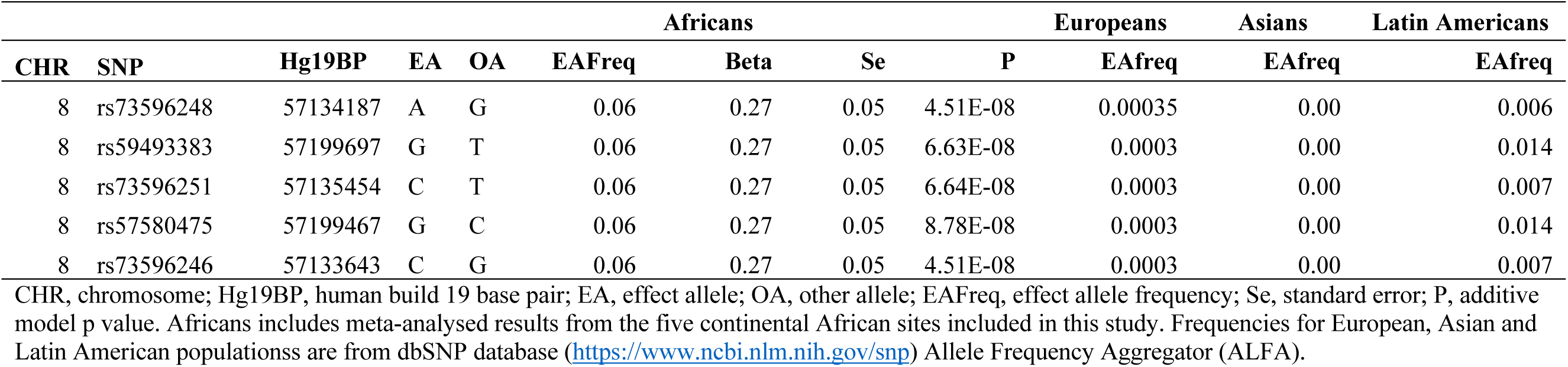
Top SNPs associated with hepcidin levels in continental African populations and their frequencies in European, Asian and Latin American populations.

**Supplementary Table 7.**
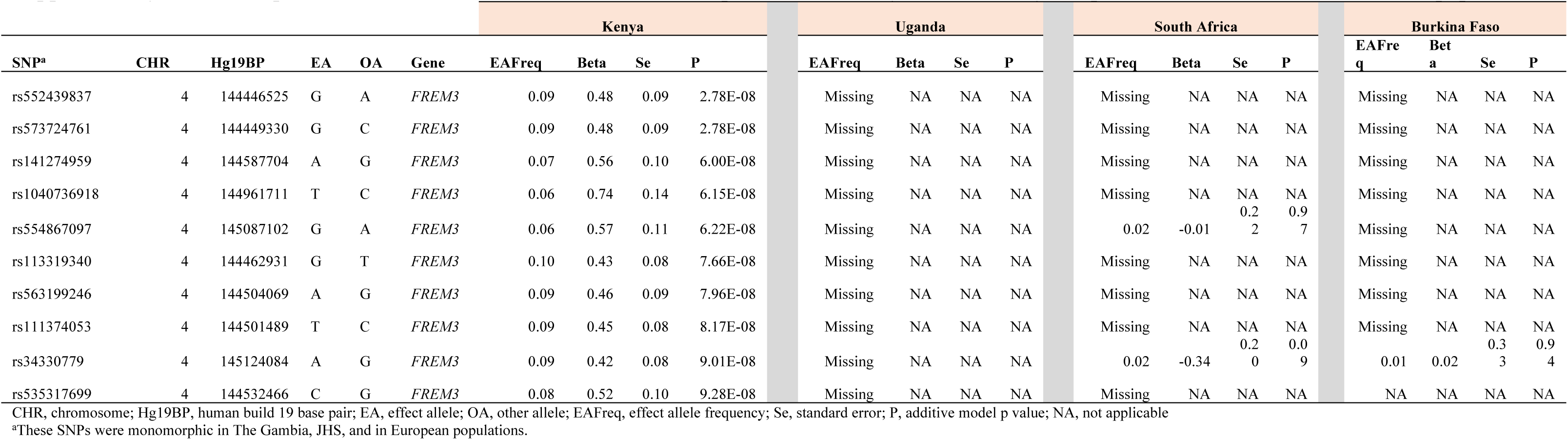
Top SNPs associated with soluble transferrin receptor levels in Kenya and how they compare with other continental African populations.

**Supplementary Table 8.** Replication of European ancestry lead hepcidin and sTfR GWAS SNPs in current study of African children.

| Trait | SNP | CHR:Hg19BP | Gene | EA | OA | European ancestry |  |  |  |  | Continental Africa |  |  |  |  |
| --- | --- | --- | --- | --- | --- | --- | --- | --- | --- | --- | --- | --- | --- | --- | --- |
|  |  |  |  |  |  | N | EAFreq | Beta | Se | P | N | EAFreq | Beta | Se | P |
| Hepcidin | rs75965181 | 1:22584002 | *MIR4418* | A | T | 91675 | 0.023 | -0.118 | 0.017 | 2.76E-12 | n/a | n/a | n/a | n/a | n/a |
| Hepcidin | rs201255457 | 1:220291414 | *IARS2* | A | ATC | 75972 | 0.989 | -0.156 | 0.026 | 3.15E-09 | n/a | n/a | n/a | n/a | n/a |
| Hepcidin | rs6706968 | 2:121310269 | *AC073257.1* | A | C | 82690 | 0.424 | 0.03 | 0.005 | 1.43E-08 | 3756 | 0.115 | 0.056 | 0.037 | 0.131 |
| Hepcidin | rs149682241 | 2:190390963 | *SLC40A1* | C | G | 91675 | 0.026 | 0.101 | 0.015 | 4.33E-11 | n/a | 0.011 | n/a | n/a | n/a |
| Hepcidin | rs7568449 | 2:190521054 | *ASNSD1* | T | C | 89099 | 0.762 | 0.043 | 0.006 | 1.88E-13 | 3756 | 0.325 | 0.017 | 0.027 | 0.525 |
| Hepcidin | rs885122 | 5:115331335 | *LVRN* | A | G | 91675 | 0.598 | -0.043 | 0.005 | 2.28E-18 | 3756 | 0.65 | -0.005 | 0.024 | 0.844 |
| Hepcidin | rs79220007 | 6:26098474 | *HFE* | T | C | 91675 | 0.93 | 0.094 | 0.01 | 1.12E-22 | n/a | n/a | n/a | n/a | n/a |
| Hepcidin | rs10090558 | 8:10577987 | *SOX7* | A | G | 78548 | 0.918 | -0.06 | 0.01 | 4.64E-10 | 3756 | 0.761 | -0.043 | 0.028 | 0.129 |
| Hepcidin | rs9739943 | 12:51439858 | *LETMD1* | T | C | 91675 | 0.059 | 0.079 | 0.01 | 3.78E-14 | 3756 | 0.201 | -0.005 | 0.029 | 0.854 |
| Hepcidin | rs11609805 | 12:57735045 | *R3HDM2* | A | G | 91675 | 0.27 | -0.032 | 0.006 | 5.24E-09 | 3756 | 0.066 | 0.041 | 0.054 | 0.45 |
| Hepcidin | rs996347 | 14:34410892 | *EGLN3* | T | C | 91675 | 0.645 | -0.033 | 0.005 | 6.06E-11 | 3756 | 0.849 | 0.012 | 0.033 | 0.718 |
| Hepcidin | rs549876436 | 15:45274530 | *TERB2* | T | C | 83099 | 0.907 | 0.082 | 0.011 | 7.30E-15 | n/a | n/a | n/a | n/a | n/a |
| Hepcidin | rs2600895 | 15:45319959 | *SORD* | A | G | 88174 | 0.913 | 0.082 | 0.012 | 2.73E-11 | n/a | n/a | n/a | n/a | n/a |
| Hepcidin | rs113000093 | 15:45320493 | *SORD* | C | G | 85598 | 0.089 | -0.078 | 0.013 | 7.01E-10 | n/a | n/a | n/a | n/a | n/a |
| Hepcidin | rs199138 | 15:45387550 | *DUOX2* | A | G | 91675 | 0.076 | -0.122 | 0.009 | 1.55E-40 | 3756 | 0.129 | 0 | 0.033 | 0.996 |
| Hepcidin | rs71378510 | 16:353122 | *AXIN1* | A | G | 91675 | 0.907 | 0.05 | 0.009 | 1.62E-08 | n/a | n/a | n/a | n/a | n/a |
| Hepcidin | rs34523089 | 17:56436109 | *RNF43* | T | C | 89817 | 0.162 | 0.063 | 0.007 | 8.15E-21 | n/a | n/a | n/a | n/a | n/a |
| Hepcidin | rs104894696 | 19:35775902 | *HAMP* | A | G | 76690 | 0.003 | -0.387 | 0.054 | 4.90E-13 | n/a | n/a | n/a | n/a | n/a |
| Hepcidin | rs485073 | 19:49207255 | *FUT2* | A | G | 91675 | 0.453 | 0.046 | 0.005 | 4.64E-21 | 3756 | 0.504 | 0.026 | 0.023 | 0.261 |
| Hepcidin | rs855791 | 22:37462936 | *TMPRSS6* | A | G | 91675 | 0.429 | 0.042 | 0.005 | 1.34E-17 | 3756 | 0.077 | 0.039 | 0.041 | 0.347 |
| sTfR | rs6696846 | 1:205041952 | *CNTN2* | T | C | 45330 | 0.484 | -0.043 | 0.007 | 6.22E-11 | 3839 | 0.214 | -0.005 | 0.027 | 0.851 |
| sTfR | rs35095338 | 2:191357694 | *TMEM194B* | A | T | 44341 | 0.556 | -0.046 | 0.007 | 9.67E-12 | 3839 | 0.554 | 0.029 | 0.025 | 0.245 |
| sTfR | rs715 | 2:211543055 | *CPS1* | T | C | 45330 | 0.688 | -0.041 | 0.007 | 2.28E-08 | 3839 | 0.828 | 0.011 | 0.032 | 0.746 |
| sTfR | rs11371594 | 3:133539500 | *SRPRB* | G | GA | 40837 | 0.845 | 0.058 | 0.01 | 2.54E-09 | n/a | n/a | n/a | n/a | n/a |
| sTfR | rs112856048 | 3:195795618 | *TFRC* | A | G | 45330 | 0.243 | -0.1 | 0.008 | 5.93E-38 | 3839 | 0.09 | 0.058 | 0.046 | 0.209 |
| sTfR | rs9325434 | 3:195921311 | *ZDHHC19* | A | G | 45330 | 0.141 | -0.062 | 0.01 | 2.27E-10 | 3839 | 0.016 | 0.115 | 0.151 | 0.444 |
| sTfR | rs2290846 | 4:151199080 | *LRBA* | A | G | 45330 | 0.277 | -0.041 | 0.007 | 3.11E-08 | n/a | n/a | n/a | n/a | n/a |
| sTfR | rs12521868 | 5:131784393 | *C5orf56* | T | G | 45330 | 0.427 | 0.052 | 0.007 | 4.41E-15 | 3839 | 0.029 | 0 | 0.074 | 0.999 |
| sTfR | rs116816795 | 5:141482333 | *NDFIP1* | T | C | 45330 | 0.168 | 0.127 | 0.009 | 3.13E-46 | 3839 | 0.028 | 0.005 | 0.105 | 0.965 |
| sTfR | rs2906082 | 5:141602204 | *NDFIP1* | T | C | 44584 | 0.783 | 0.057 | 0.008 | 5.34E-12 | 3839 | 0.447 | 0.034 | 0.023 | 0.133 |
| sTfR | rs72832593 | 6:25957426 | *TRIM38* | T | C | 45330 | 0.897 | 0.13 | 0.011 | 8.09E-33 | n/a | n/a | n/a | n/a | n/a |
| sTfR | rs1800562 | 6:26093141 | *HFE* | A | G | 45330 | 0.075 | -0.253 | 0.013 | 2.88E-88 | n/a | n/a | n/a | n/a | n/a |
| sTfR | rs9389269 | 6:135427159 | *HBS1L* | T | C | 45330 | 0.729 | 0.059 | 0.007 | 3.10E-15 | 3839 | 0.961 | -0.023 | 0.078 | 0.77 |
| sTfR | rs143437464 | 6:159020121 | *TMEM181* | A | G | 43595 | 0.009 | -0.234 | 0.037 | 3.81E-10 | n/a | n/a | n/a | n/a | n/a |
| sTfR | rs200307986 | 6:159026327 | *TMEM181* | A | G | 42671 | 0.003 | 0.358 | 0.065 | 2.69E-08 | n/a | n/a | n/a | n/a | n/a |
| sTfR | rs186044114 | 7:1080897 | *C7orf50* | T | C | 41188 | 0.852 | -0.061 | 0.01 | 4.19E-10 | n/a | n/a | n/a | n/a | n/a |
| sTfR | rs6592965 | 7:50427982 | *IKZF1* | A | G | 45330 | 0.458 | -0.058 | 0.007 | 2.43E-18 | 3839 | 0.501 | 0.013 | 0.023 | 0.575 |
| sTfR | rs7385804 | 7:100235970 | *TFR2* | A | C | 45330 | 0.627 | -0.051 | 0.007 | 1.49E-13 | 3839 | 0.726 | 0 | 0.026 | 0.988 |
| sTfR | rs7908745 | 10:45953767 | *MARCH8* | A | G | 45330 | 0.687 | -0.054 | 0.007 | 5.61E-14 | 3839 | 0.677 | 0.027 | 0.024 | 0.262 |
| sTfR | rs16926246 | 10:71093392 | *HK1* | T | C | 45330 | 0.133 | 0.056 | 0.01 | 1.05E-08 | 3839 | 0.164 | -0.045 | 0.031 | 0.143 |
| sTfR | rs187669805 | 11:117021097 | *PCSK7* | C | G | 43487 | 0.994 | 0.407 | 0.046 | 1.94E-18 | n/a | n/a | n/a | n/a | n/a |
| sTfR | rs11216316 | 11:117081500 | *PCSK7* | A | C | 45330 | 0.892 | -0.273 | 0.011 | 1.77E-137 | 3839 | 0.83 | -0.041 | 0.032 | 0.196 |
| sTfR | rs2238005 | 11:117088082 | *PCSK7* | T | C | 45330 | 0.061 | 0.118 | 0.014 | 2.71E-17 | 3839 | 0.031 | -0.066 | 0.086 | 0.441 |
| sTfR | rs10876169 | 12:51783420 | *GALNT6* | A | T | 44584 | 0.41 | 0.039 | 0.007 | 1.85E-08 | 3839 | 0.032 | -0.005 | 0.081 | 0.955 |
| sTfR | rs76944188 | 13:110401304 | *IRS2* | T | C | 45330 | 0.942 | -0.09 | 0.015 | 5.45E-10 | 3839 | 0.794 | 0.006 | 0.028 | 0.818 |
| sTfR | rs75922593 | 15:45395901 | *DUOX2* | G | GT | 40837 | 0.066 | 0.112 | 0.014 | 3.64E-15 | n/a | n/a | n/a | n/a | n/a |
| sTfR | rs74035509 | 16:88567333 | *ZFPM1* | T | C | 44584 | 0.083 | 0.083 | 0.013 | 6.36E-11 | 3839 | 0.15 | -0.02 | 0.033 | 0.551 |
| sTfR | rs55925547 | 17:43556807 | *PLEKHM1* | T | C | 43244 | 0.801 | -0.057 | 0.009 | 2.46E-11 | 3839 | 0.837 | 0.034 | 0.031 | 0.276 |
| sTfR | rs1292072 | 17:57925649 | *RNU6-450P* | A | G | 45330 | 0.794 | -0.048 | 0.008 | 7.13E-09 | 3839 | 0.945 | -0.01 | 0.048 | 0.828 |
| sTfR | rs1976703 | 17:76401328 | *PGS1* | T | C | 41826 | 0.493 | 0.039 | 0.007 | 1.57E-08 | 3839 | 0.3 | -0.006 | 0.025 | 0.793 |
| sTfR | rs13041 | 19:4502282 | *PLIN4* | T | C | 44584 | 0.518 | 0.048 | 0.007 | 8.53E-12 | 3839 | 0.332 | -0.014 | 0.025 | 0.591 |
| sTfR | rs855791 | 22:37462936 | *TMPRSS6* | A | G | 45330 | 0.434 | 0.134 | 0.007 | 2.70E-89 | 3839 | 0.077 | 0.001 | 0.041 | 0.973 |

European ancestry summary statistics were obtained from the most recent and largest GWAS of hepcidin and sTfR by Allara et al 2024.
CHR:BP(hg19), chromosome and human genome build 19 base pair; EA, effect allele; OA, other allele; EAFreq, effect allele frequency; Se, standard error; P, additive model p value; n/a, not available

**Supplementary Table 9.**
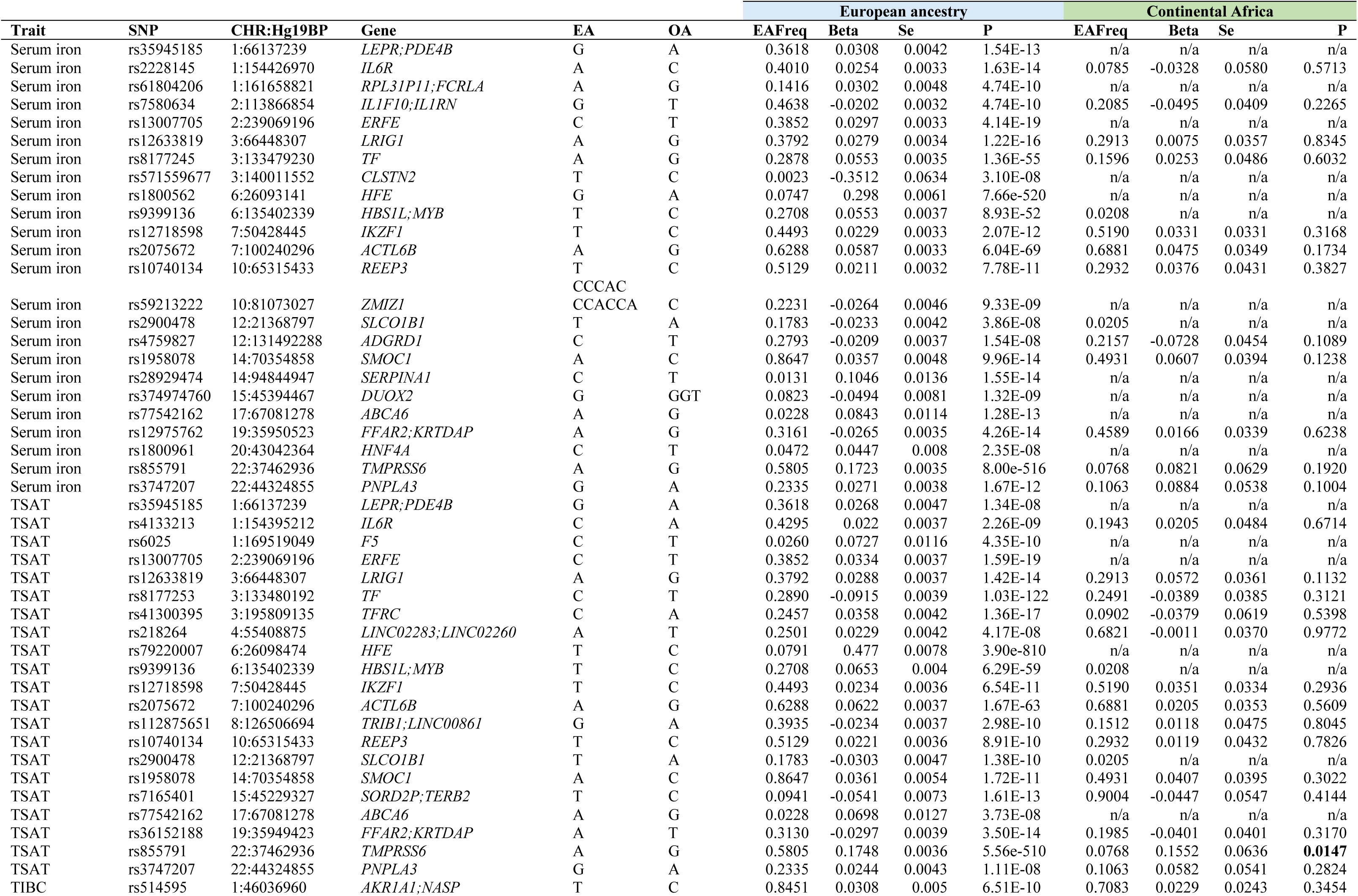
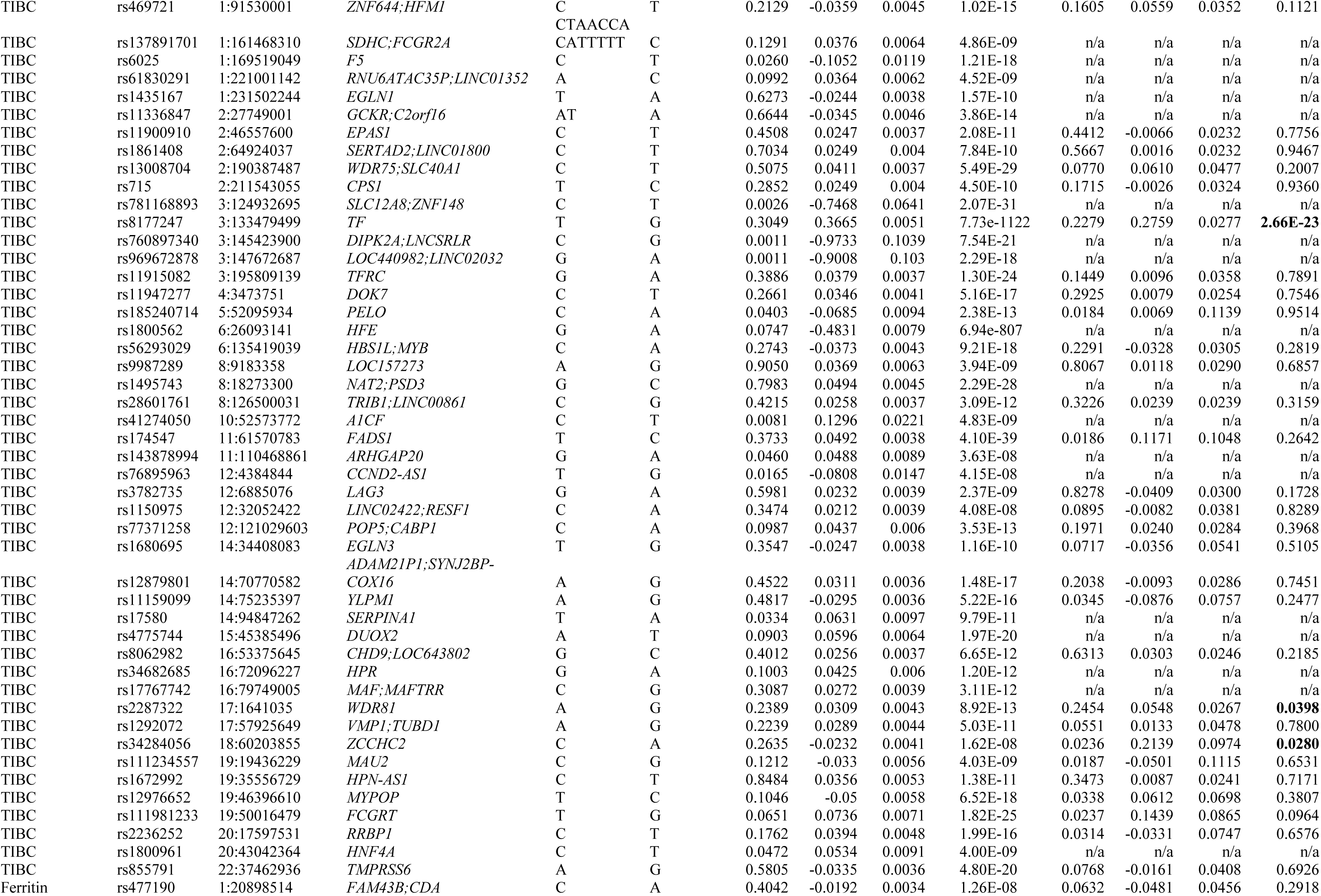
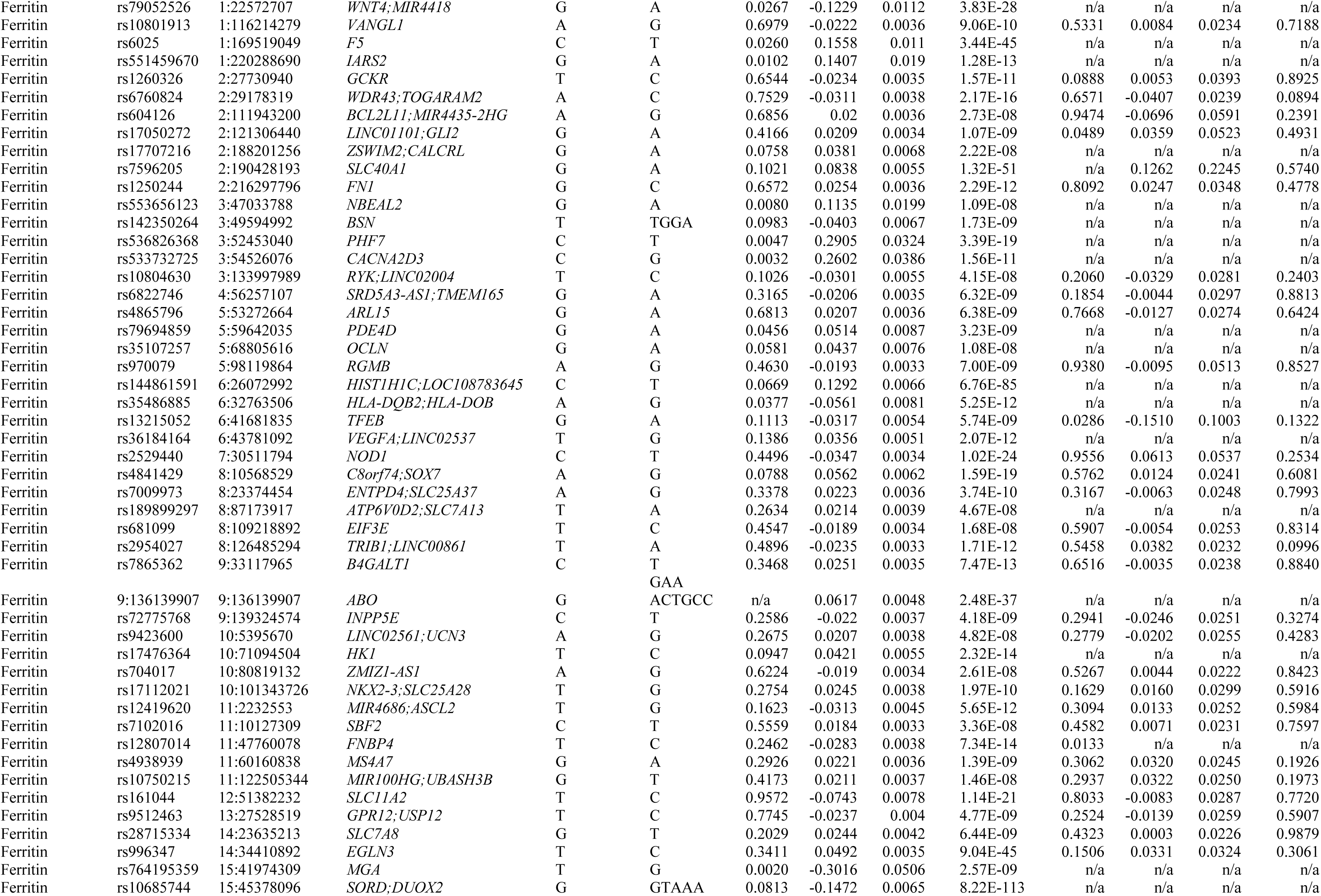
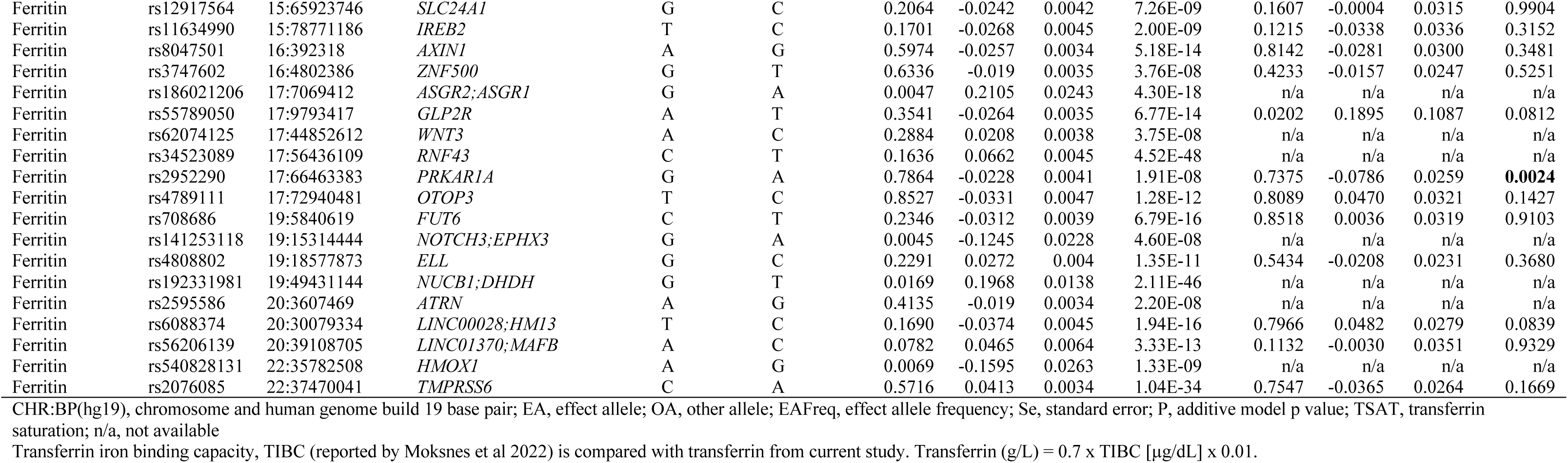
Replication of European ancestry lead serum iron, transferrin saturation, TIBC and ferritin GWAS SNPs in current study of African children.

**Supplementary Table 10.** Laboratory assays for iron and inflammatory biomarkers by site.

| Study site | Transferrin | Ferritin | sTfR | Hepcidin | Iron | UIBC | TIBC | CRP | ACT |
| --- | --- | --- | --- | --- | --- | --- | --- | --- | --- |
| Kilfi, Kenya | Chemiluminescent Microparticle Immunoassay (Abbott Architect, USA) | Microparticle Enzyme Immunoassay (Abbott Architect, USA) | Human sTfR ELISA (BioVendor, Czech Republic) | DRG Hepcidin 25 [bioactive] high sensitive ELISA (DRG International, USA) | MULTIGENT iron calorimetric assay, Abbott Architect, USA | Not analyzed | Not analyzed | MULTIGENT CRP Vario assay, (Abbott Architect, USA) | Not analyzed |
| Banfora,Burkina Faso | Chemiluminescent Microparticle Immunoassay (Abbott Architect, USA) | Microparticle Enzyme Immunoassay (Abbott Architect, USA) | Human sTfR ELISA (BioVendor, Czech Republic) | DRG Hepcidin 25 [bioactive] high sensitive ELISA (DRG International, USA) | MULTIGENT iron calorimetric assay, Abbott Architect, USA | Not analyzed | Not analyzed | MULTIGENT CRP Vario assay, (Abbott Architect, USA) | Not analyzed |
| Entebbe, Uganda | Chemiluminescent Microparticle Immunoassay (Abbott Architect, USA) | Microparticle Enzyme Immunoassay (Abbott Architect, USA) | Human sTfR ELISA (BioVendor, Czech Republic) | DRG Hepcidin 25 [bioactive] high sensitive ELISA (DRG International, USA) | Not analyzed | Not analyzed | Not analyzed | MULTIGENT CRP Vario assay, (Abbott Architect, USA) | Not analyzed |
| Soweto, South Africa | Chemiluminescent Microparticle Immunoassay (Abbott Architect, USA) | Microparticle Enzyme Immunoassay (Abbott Architect, USA) | Human sTfR ELISA (BioVendor, Czech Republic) | DRG Hepcidin 25 [bioactive] high sensitive ELISA (DRG International, USA) | Not analyzed | Not analyzed | Not analyzed | MULTIGENT CRP Vario assay, (Abbott Architect, USA) | Not analyzed |
| West Kiang, The Gambia | Not analyzed | Microparticle Enzyme Immunoassay (Abbott Architect, USA) | Quantikine sTfR ELISA kit, (R&D Systems, USA) | Hepcidin-25 [human] Enzyme Immunoassay Kit (Bachem, Switzerland) | Ferrozine-based photometry and colorimetry analyser (Hitachi 911, Hitachi, Tokyo, Japan) | Ferrozine-based photometry and colorimetry analyser (Hitachi 911, Hitachi, Tokyo, Japan) | Not analyzed | Not analyzed | Immunoturbidimetry, Cobas Mira Plus Bio-analyser, Roche |
| Jackson Heart Study | Not analyzed | Roche immunoturbidimetric assay | Not analyzed | Not analyzed | Ferrozine colorimetric assay (Roche) | Not analyzed | Ferrozine colorimetric assay (Roche) | Immunoturbidimetric CRP-Latex assay (Roche) | Not analyzed |

sTfR, soluble transferrin receptor; UIBC, unsaturated iron binding capacity; TIBC, total iron binding capacity; CRP, C-reactive protein; ACT, α1-antichymotrypsin

